# Machine learning for predicting severe dengue, Puerto Rico

**DOI:** 10.1101/2024.11.15.24317377

**Authors:** Zachary J. Madewell, Dania M. Rodriguez, Maile B. Thayer, Vanessa Rivera-Amill, Gabriela Paz-Bailey, Laura E. Adams, Joshua M. Wong

## Abstract

**Background:** Distinguishing between non-severe and severe dengue is crucial for timely intervention and reducing morbidity and mortality. Traditional warning signs recommended by the World Health Organization (WHO) offer a practical approach for clinicians but have limitations in sensitivity and specificity. This study evaluates the performance of machine learning (ML) models compared to WHO-recommended warning signs in predicting severe dengue among laboratory-confirmed cases in Puerto Rico.

**Methods:** We analyzed data from Puerto Rico’s Sentinel Enhanced Dengue Surveillance System (May 2012–August 2024), using 40 clinical, demographic, and laboratory variables. Nine ML models, including Decision Trees, K-Nearest Neighbors, Naïve Bayes, Support Vector Machines, Artificial Neural Networks, AdaBoost, CatBoost, LightGBM, and XGBoost, were trained using 5-fold cross-validation and evaluated with area under the receiver operating characteristic curve (AUC-ROC), sensitivity, specificity, positive predictive value (PPV), and negative predictive value (NPV). A subanalysis excluded hemoconcentration and leukopenia to assess performance in resource-limited settings. An AUC-ROC value of 0.5 indicates no discriminative power, while a value closer to 1.0 reflects better performance.

**Results:** Among the 1,708 laboratory-confirmed dengue cases, 24.3% were classified as severe. Gradient boosting algorithms achieved the highest predictive performance, with AUC-ROC values exceeding 94% for CatBoost, LightGBM, and XGBoost. Feature importance analysis identified hemoconcentration (≥20% increase during illness or ≥20% above baseline for age and sex), leukopenia (white blood cell count <4,000/mm³), and timing of presentation to a healthcare facility at 4–6 days post-symptom onset as key predictors. Excluding hemoconcentration and leukopenia did not significantly affect model performance. Individual warning signs like abdominal pain and restlessness had sensitivities of 79.0% and 64.6%, but lower specificities of 48.4% and 59.1%, respectively. Combining ≥3 warning signs improved specificity (80.9%) while maintaining moderate sensitivity (78.6%), resulting in an AUC-ROC of 74.0%.

**Conclusions:** ML models, especially gradient boosting algorithms, outperformed traditional warning signs in predicting severe dengue. Integrating these models into clinical decision-support tools could help clinicians better identify high-risk patients, guiding timely interventions like hospitalization, closer monitoring, or the administration of intravenous fluids. The subanalysis excluding hemoconcentration confirmed the models’ applicability in resource-limited settings, where access to laboratory data may be limited.

## Background

Dengue is a significant public health concern worldwide, with approximately 390 million infections annually, of which 96 million manifest clinically [1, 2]. In Puerto Rico, dengue has been associated with nearly 30,000 confirmed and probable cases from 2010 to 2020, including 584 severe cases, 10,000 hospitalizations, and 68 deaths [3]. A surge in dengue cases on the island in 2024 prompted a public health emergency declaration by Puerto Rico’s Department of Health, highlighting the ongoing threat of dengue to the island [4]. These regular outbreaks strain healthcare resources and pose substantial morbidity and mortality risks. A critical aspect of managing dengue is distinguishing between non-severe and severe cases, as the latter require intensive medical intervention to prevent complications and fatalities. Early identification of patients at risk of severe dengue is important for timely intervention and improved patient outcomes. However, predicting which patients will progress to severe dengue remains a challenge, often leading to delayed treatment and increased healthcare burden.

The World Health Organization (WHO) has recommended identifying severe dengue through clinical assessment of warning signs such as persistent vomiting, abdominal pain, mucosal bleeding, restlessness, and hepatomegaly [5]. Although these warning signs offer a practical approach for clinicians, their specificity and sensitivity in accurately predicting severe dengue are limited. Studies have shown that relying solely on these warning signs can result in both false positives and negatives, potentially leading to over- or under-treatment of patients [6–10]. The substantial burden of dengue on the healthcare system, both in terms of economic cost and human suffering, underscores the need for innovative approaches to disease diagnosis. A more accurate and efficient method for risk stratification could lead to substantial improvements in patient care and resource allocation.

In recent years, machine learning (ML) has emerged as a powerful tool for analyzing complex datasets and uncovering patterns not easily discernible by traditional methods. In the context of dengue, ML models can analyze a multitude of factors beyond the established warning signs, including patient demographics, laboratory results, clinical symptoms, and epidemiological data, to enhance the prediction of severe disease [11, 12]. By leveraging ML, we aim to improve the accuracy of severe dengue predictions, offering a more robust and data-driven approach to risk stratification. If successful, these models could transform dengue management by enabling early, accurate identification of high-risk patients, ultimately leading to improved patient outcomes and reduced mortality rates. Additionally, targeted intervention for high-risk patients can optimize resource allocation, ensuring critical care is available to those who need it most.

This project specifically leverages data from Puerto Rico’s Sentinel Enhanced Dengue Surveillance System (SEDSS), which has detailed clinical and laboratory information on dengue cases, allowing for the exploration of potential predictors of disease severity beyond the conventional warning signs. In addition to exploring ML approaches, this project also aims to evaluate the performance of WHO-recommended warning signs in predicting severe dengue among laboratory-confirmed cases, which include both molecular (RT-PCR) and serologic (IgM ELISA) testing. By doing so, we seek to bridge the gap between research and clinical practice by demonstrating the practical applications of both traditional and advanced computational tools in identifying severe dengue. If our findings demonstrate that ML models offer improved prediction of severe dengue compared to traditional methods, this could highlight the potential for integrating advanced computational tools into public health strategies and clinical protocols. For example, predictive ML models could be incorporated into clinical decision support systems used in emergency departments or outpatient clinics, enabling real-time risk stratification for severe dengue. This integration could help healthcare providers prioritize patients for hospitalization, allocate medical resources more efficiently, and guide timely interventions to prevent complications and fatalities. These tools could enable early and accurate identification of high-risk patients, improve patient outcomes, and optimize resource allocation.

## Methods

### Study population

In this analysis, we used data from SEDSS, an ongoing facility-based study in Puerto Rico that tracks the frequency and causes of acute febrile illness [13, 14]. Our study included data from SEDSS from May 2012 to August 2024. SEDSS has included five sites: 1) Centro Médico Episcopal San Lucas (CMESL) in Ponce, a tertiary acute care facility (2012-present), 2) Hospital Episcopal San Lucas (HESL) - Guayama, a secondary acute care hospital (2013–2015), 3) Hospital de La Universidad de Puerto Rico in Carolina, another secondary acute care teaching hospital (2013–2015), 4) Centro de Emergencia y Medicina Integrada (CEMI), an outpatient acute care clinic in Ponce (2016-present), and 5) Auxilio Mutuo Hospital, a tertiary care facility in the San Juan Metro Area (2018–present).

### Study enrollment and data collection

SEDSS enrolls participants using convenience sampling. Potential participants are identified by triage nurses as any patient with an acute febrile illness (AFI) defined by the presence of fever (≥38.0°C for temperatures measured orally, ≥37.5°C for temperatures measured rectally, and ≥38.5°C for temperatures measured axillarily for both children and adults) at the time of triage or chief complaint of having a fever within the past seven days. During the Zika virus epidemic in Puerto Rico (June 2016– June 2018), patients were eligible if they presented with either rash and conjunctivitis, rash and arthralgia, or fever [15]. Starting in April 2020, patients with cough or dyspnea within the last 14 days (with or without fever) were also eligible to better capture respiratory viruses [16]. No age groups were excluded, although infants were only eligible for enrollment if they presented to the hospital after their initial discharge after birth. After meeting the inclusion criteria and being informed about the study, participants provided written informed consent. In cases where patients were incapacitated at the time of triage due to acute illness, consent was sought after their stabilization.

SEDSS collects data via patient interviews and medical record reviews at enrollment and convalescence (∼7–14 days later). The case investigation form (CIF) gathers information about patient demographics, comorbidities, and clinical features. The convalescent sample processing form (CSPF) echoes CIF data, adding the second specimen collection date and AFI severity indicators (hospitalizations, clinic visits). Inpatient medical data for participants with AFIs who were admitted to the hospital from CMESL, HESL-Guayama, and Auxilio Mutuo Hospital also were collected using a separate form (Hospital Admitted Abstraction Form) to collect key clinical indicators of disease severity and progression. For admitted patients, these data included information on extent and nature of hemorrhage, plasma leakage (e.g., ascites and pleural and cardiac effusions), hematologic indicators of increased intravascular permeability (e.g., hematocrit and serum albumin levels), additional blood pressure and heart rate measures to assess shock, and indicators of severe organ involvement (e.g., liver impairment, meningitis, and encephalitis) [14].

Dengue warning signs and severe dengue were defined by the World Health Organization [17], incorporating available clinical indicators from SEDSS intake and follow-up forms and abstracted inpatient medical records. Dengue warning signs were defined by abdominal pain or tenderness, persistent vomiting, plasma leakage (pleural or pericardial effusion or ascites), mucosal bleeding, restlessness, hemoconcentration (defined as either a hematocrit increase of ≥20% during illness or a hematocrit value ≥20% above baseline for age and sex), or hepatomegaly. Detailed definitions for these variables have been provided previously [18]. Severe dengue was defined as severe plasma leakage or shock, severe bleeding, or severe organ impairment [18]. The presence and overlap of warning signs among severe dengue cases were visualized using an Euler plot via the eulerr R package [19].

### Sample collection and laboratory procedures

Blood, nasopharyngeal (NP), and oropharyngeal (OP) specimens were collected at enrollment from eligible participants. Additional blood samples (serum and whole blood) were also collected during the convalescent phase. Participation required providing at least one sample (blood or OP/NP swab). All patients had molecular testing for dengue virus for specimens collected within 7 days of symptom onset. Serologic testing was done by Immunoglobulin M (IgM) antibody capture enzyme-linked immunosorbent assay (ELISA) for anti-DENV antibodies for specimens collected >3 days after symptom onset [20].

### Variables

A total of 40 variables were selected based on the WHO’s criteria for severe dengue, physicians’ clinical experience, and a review of current literature to potentially differentiate between severe and non-severe dengue cases [17, 18, 21–24]. These features included age group, days post onset of symptoms, clinical signs and symptoms, laboratory findings, pre-existing health conditions, and dengue virus serotype. Clinical symptoms included report of fever, rash, headache, myalgia, abdominal pain, chills, itchy skin, eye pain, nasal discharge, cough, sore throat, persistent vomiting, diarrhea, arthralgia, arthritis, back pain, calf pain, nausea, no appetite, and restlessness. Clinical signs, as observed by healthcare providers, included objective fever at the time of enrollment, yellow skin (jaundice), observed bruising, conjunctivitis, hepatomegaly, mucosal bleeding, pale skin, and blue lips (cyanosis). Clinical laboratory findings comprised leukopenia (defined as white blood cell count <4,000/mm³), the calculated value of hemoconcentration (an increase in the concentration of red blood cells due to plasma loss), and dengue immune status (primary or post-primary) as measured from results for anti-dengue virus immunoglobulin G (IgG) on or before day 5 of illness. Pre-existing health conditions like obesity (BMI ≥ 30), gastritis, chronic arthritis, hypertension, chronic kidney disease, diabetes, thyroid disease, and high cholesterol were also considered. Additionally, dengue virus serotype was included, coded as “unknown” for probable cases identified through IgM ELISA, as serotype data was only available for confirmed reverse transcription polymerase chain reaction (RT-PCR) cases. Thrombocytopenia and clinical fluid accumulation were excluded from the analysis, as they generally manifest after the onset of severe disease or are components of its definition, making them less useful as predictive features. This comprehensive set of variables was intended to represent the multifaceted nature of factors influencing dengue infection severity.

### Sensitivity analyses

To further explore the performance of the models in resource-constrained settings, where complete blood counts (CBCs), dengue immune status, and serotype information might not be readily available, we performed sub-analyses to evaluate model robustness and applicability. The sub-analyses included: 1) excluding results found on a CBC (i.e., leukopenia and hemoconcentration), 2) excluding IgG and serotype results, and 3) excluding leukopenia, hemoconcentration, IgG, and serotype results. These analyses were designed to assess how well the models could predict progression to severe dengue in various clinical scenarios, particularly where access to comprehensive clinical laboratory results or pathogen-specific testing might be limited.

To ensure the robustness of our models and account for potential confounding from co-circulating arboviruses, we performed a sensitivity analysis with the highest-performing individual ML model, excluding cases that tested positive for chikungunya virus (CHIKV) by either IgM or RT-PCR. This analysis aimed to confirm that the predictive features for severe dengue remain consistent even in the absence of CHIKV, given the potential overlap in clinical presentations between the two viruses.

### Sampling

Our analysis included laboratory-confirmed dengue cases, confirmed by either molecular or serologic testing, focusing on differentiating between severe and non-severe cases. Due to an imbalance in the dataset, where non-severe cases were more prevalent, we used upsampling to balance the class distribution. Upsampling involves increasing the number of minority class samples (severe dengue cases) through duplication, which helps to prevent model bias towards the majority class and improves the model’s ability to accurately predict severe dengue cases [25]. Upsampling was done using the upSample function from the caret package in R [26]. Following upsampling, the dataset was divided into training and testing sets using a 70/30 split: 70% of the data was allocated for training the models, while the remaining 30% was reserved for testing. This partitioning ensured that the models were trained on a substantial portion of the data while retaining a sufficient amount for unbiased evaluation.

### Machine learning models

An initial logistic regression (LR) model served as a baseline simple model to explore the relationship between potential predictors and the outcome of severe dengue. Stepwise selection, implemented using the stepAIC function from the MASS package in R [27], was used to iteratively add or remove variables to identify the optimal model with the lowest Akaike Information Criterion. This approach balances model complexity and goodness-of-fit by selecting variables that contribute significantly to the model. The final logistic regression model, derived from stepwise selection, was evaluated on both the training and testing sets.

In addition, we used nine ML methods to predict severe dengue and analyze feature importance. A comprehensive selection of models was used to leverage different strengths, enhance predictive performance, and provide a nuanced understanding of the factors contributing to severe dengue. The algorithms used include Decision Trees (DT), K-Nearest Neighbors (KNN), Naïve Bayes, Support Vector Machines (SVM), Artificial Neural Networks (ANN), Adaptive Boosting (AdaBoost), Categorical Boosting (CatBoost), Light Gradient Boosting Machine (LightGBM), and eXtreme Gradient Boosting (XGBoost). DTs create a tree-like structure to make predictions by recursively splitting the data based on feature values [28]. KNNs predict the class of a data point by considering the majority class of its closest neighbors in the feature space [29]. Naïve Bayes is a probabilistic classifier that applies Bayes’ theorem, assuming independence between predictors [30]. SVMs identify the optimal hyperplane to separate classes, making them effective for high-dimensional data [31]. ANNs are inspired by biological neural networks and consist of interconnected nodes that can capture complex patterns [32]. AdaBoost, CatBoost, LightGBM, and XGBoost are ensemble methods that combine multiple weak learners to improve predictive performance [33, 34]. AdaBoost adjusts weights to focus on difficult-to-predict instances, CatBoost handles categorical features effectively, LightGBM is efficient with large datasets due to its leaf-wise tree growth, and XGBoost uses regularization techniques to prevent overfitting, enhancing accuracy and robustness [33–35].

The hyperparameters (model-specific settings, such as learning rate, maximum tree depth, or number of estimators) for each ML model were carefully tuned using a grid search strategy to optimize performance. The area under the receiver operating characteristic curve (AUC-ROC) was used as the optimization metric, ensuring a focus on maximizing classification performance. We used 5-fold cross-validation during model training to enhance robustness and mitigate overfitting. Specific details of the grid search strategy and parameters included in each model are provided in Table S1.

The following R packages were used for model implementation: rpart [36] for DT, e1071 [37] for Naïve Bayes and SVM, caret [26] for KNN, nnet [38] for ANN, keras [39] for DNN, ada [40] for AdaBoost, catboost [41] for CatBoost, lightgbm [42] for LightGBM, and xgboost [43] for XGBoost. All analyses were done using R version 4.4.0 [44].

### Ensemble model

To leverage the predictive power of multiple ML algorithms, we used an ensemble learning approach with a stacked generalization framework. This method combines the strengths of various individual models to improve overall predictive performance and robustness. We used predictions from LR and the nine different ML models as base learners in our ensemble. Specifically, we used a logistic regression model as the meta-learner to combine the outputs of the base models. This approach allows the meta-model to learn the optimal combination of base models’ predictions. To improve the performance of the meta-model, we again used stepwise selection with the stepAIC function from the MASS package [27].

Pearson correlation coefficients were calculated to measure the linear correlation between the predictions of the ML models. This analysis helps determine whether the models are making similar predictions for severe dengue, potentially reflecting the selection of similar variables and patterns across the models. The results were visualized in a heatmap using ggplot2 [45].

### Performance evaluation

Model performance for each ML model and the meta-model was evaluated on both the training and testing sets using AUC-ROC as the primary performance metric. AUC-ROC is an aggregate measure of performance across all possible classification thresholds, providing a comprehensive assessment of the model’s ability to distinguish between classes. We used the DeLong method to calculate the confidence intervals for the AUC-ROC to ensure accurate estimation of the model’s performance [46].

The performance of the ensemble model was further evaluated using several metrics, including accuracy, sensitivity, specificity, positive predictive value (PPV), negative predictive value (NPV), F1 score, and Cohen’s kappa. These metrics provide a comprehensive view of the model’s performance, capturing both the ability to correctly classify severe dengue cases and the overall agreement between predicted and actual classifications.

### Feature importance

Feature importance was calculated for each ML model to quantify the contribution of each variable to the model’s predictive accuracy. This approach enables the identification of the most influential features, which enhances our understanding of the factors driving the predictions for severe dengue. Feature importance was assessed for both the 40-variable feature set and a subset excluding CBCs, IgG, and serotype results. Different methods were applied across the ML algorithms to determine feature importance. For ensemble-based methods, including XGBoost, LightGBM, and CatBoost, feature importance was calculated using the Gain metric, which measures the contribution of each feature to the model’s decision-making process. Gain represents the improvement in the model’s accuracy brought by a feature, with higher values indicating greater importance. For XGBoost, LightGBM, and CatBoost, we used the xgb.importance, lgb.importance, and catboost.get_feature_importance functions from the xgboost [43], lightgbm [42], and catboost [41] packages, respectively.

Permutation importance was applied to assess feature importance for KNN, Naive Bayes, and ANN. This method involves randomly shuffling feature values and measuring the subsequent decline in model performance. A substantial decrease in accuracy indicates a highly influential feature. For DT, feature importance was determined by the reduction in impurity (Gini index or entropy) achieved by splitting data based on that feature. AdaBoost assigned importance to features based on their contribution to correcting errors in subsequent models, with higher weights indicating greater influence. For SVM, feature importance was derived from the absolute value of the model coefficients. The magnitude of these coefficients reflects the influence of each feature on the decision boundary, with larger coefficients indicating greater importance.

We also calculated and plotted SHapley Additive exPlanations (SHAP) values for the top three performing models based on AUC-ROC. SHAP values provide a nuanced measure of each feature’s contribution to the prediction of severe dengue cases, enabling a deeper understanding of model decision-making. Positive SHAP values indicate a higher likelihood of severe dengue, whereas negative values suggest a protective effect. The SHAP approach is particularly valuable as it allows for the decomposition of the prediction into individual feature contributions, offering a clear interpretation of how different variables influence the model’s predictions.

### Post-hoc variable reduction analysis

To assess the predictive performance of a simplified variable set, we conducted a post-hoc analysis using the ML model that achieved the highest AUC. This analysis aimed to identify the minimum number of features needed to maintain high accuracy. We began with the top features identified through SHAP values in the original 40-variable analysis, adding one feature at a time, starting with the highest-ranking. At each step, we evaluated the AUC-ROC to determine the impact of including additional features. The goal was to develop a more streamlined model that remains feasible and interpretable, especially in clinical settings with limited diagnostic resource.

### Diagnostic accuracy of warning signs

In addition to ML, we evaluated the diagnostic accuracy of individual warning signs for identifying severe dengue cases. The performance of each warning sign was assessed using sensitivity, specificity, PPV, NPV, and AUC-ROC. Sub-analyses assessed the performance of warning signs by dengue serotype and immune status. Dengue serotype was determined via RT-PCR, whereas immune status was classified based on IgG antibody results in the first 5 days after illness onset (primary: IgG; post-primary: positive IgG). Cases lacking serotype or immune status data were excluded from sub-analyses. This approach aimed to identify potential clinical differences in the presentation and predictive capacity of warning signs for severe dengue across serotypes and infection statuses.

### Ethics statement

The Institutional Review Boards at the Centers for Disease Control and Prevention (CDC), Auxilio Mutuo, and Ponce Medical School Foundation approved the SEDSS study protocols 6214, and 120308-VR/2311173707, respectively. Written consent to participate was obtained from all adult participants and emancipated minors. For minors aged 14 to 20 years, written consent was obtained, and for those aged 7 to 13 years, parental written consent and participant assent were obtained.

## Results

### Characteristics of dengue cases

From May 2012 to August 2024, there were 51,877 unique AFI visits from 41,647 participants enrolled in SEDSS, including 8,404 hospitalizations or transfers and 75 deaths. Of these visits, there were 50,189 AFI visits from 40,495 participants tested for DENV. From these, 1,708 (3.4%) had dengue (1,218 confirmed, 490 probable). The majority of the 1,206 serotyped dengue cases were DENV-1 (n=905, 75.0%), followed by DENV-3 (n=149, 12.4%), DENV-2 (n=102, 8.5%), and DENV-4 (n=50, 4.1%). Of 1,708 dengue cases, 759 (44.4%) were hospitalized or transferred, and two (0.1%) died. The median duration from symptom onset to presentation at the emergency room was 3 days [IQR: 2, 5]. Of the 730 participants assessed for immune status using DENV IgG, 577 (79.0%) were positive, indicating post-primary dengue, whereas the remaining 153 (21.0%) were negative, suggesting primary dengue infections.

Among the 1,708 laboratory-confirmed dengue cases, 24.3% (n=415) were classified as severe dengue. Compared to those without severe dengue, participants with severe dengue were more likely to present between 4 to 6 days post-symptom onset (52.4% vs. 32.0%, *p* < 0.001) and be aged 10-19 years (51.6% vs. 36.3%, *p* < 0.001) (Table 1). Among dengue cases tested, a higher proportion of severe cases were post-primary DENV infections (85.7% vs. 76.3%, *p* = 0.007). Participants with severe dengue had a higher prevalence of warning signs such as persistent vomiting (37.6% vs. 20.4%), abdominal pain (79.0% vs. 51.6%), restlessness (64.6% vs. 40.9%), mucosal bleeding (22.9% vs. 13.8%), and hemoconcentration (20.7% vs. 3.3%) compared to lab-confirmed non-severe cases (all *p* < 0.001) (Figure 1). All 30 dengue cases with seizures were classified as severe dengue. Leukopenia (77.1% vs. 53.5%) was more prevalent among participants with severe dengue (*p* < 0.001).

**Figure 1.**
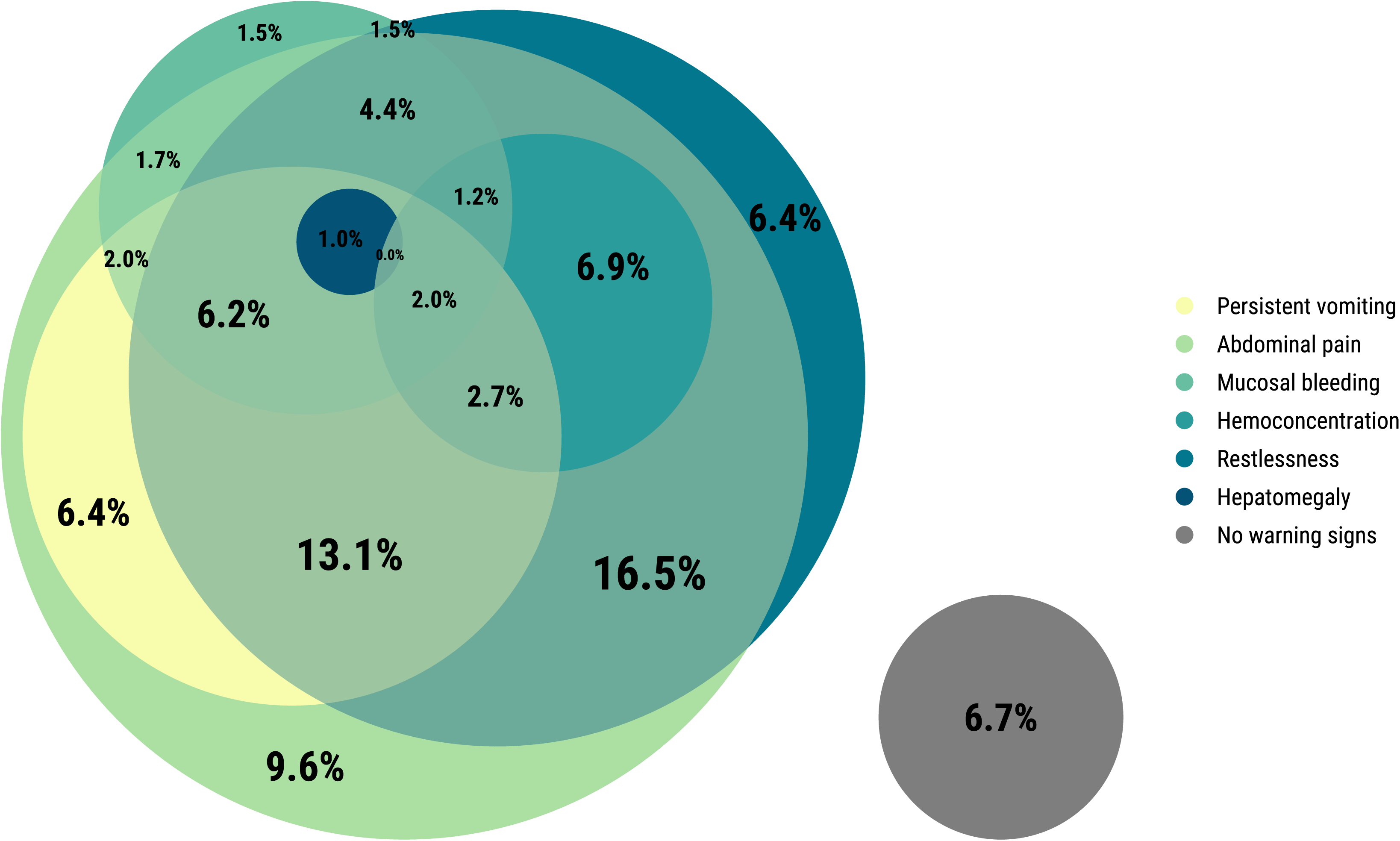
Euler plot of proportion of severe dengue cases with each warning sign, Sentinel Enhanced Dengue Surveillance System, Puerto Rico, 2012–2024.

**Table 1.** Demographic and clinical characteristics of participants with laboratory-confirmed dengue (RT-PCR and IgM ELISA) by severity, Sentinel Enhanced Dengue Surveillance System, Puerto Rico, 2012–2024.

|  | **Total**<br>**N = 1,708**<br>**n (column %)** | **Severe dengue**<br>**N = 415**<br>**n (column %)** | **Not severe**<br>**N = 1,293**<br>**n (column %)** | **p-value** |
| --- | --- | --- | --- | --- |
| **Days post onset** |  |  |  | <0.001 |
| 0 | 128 (7.5) | 18 (4.3) | 110 (8.5) |  |
| 1-3 | 810 (47.4) | 133 (32.0) | 677 (52.4) |  |
| 4-6 | 695 (40.7) | 243 (58.6) | 452 (35.0) |  |
| 7+ | 75 (4.4) | 21 (5.1) | 54 (4.2) |  |
| **Age group** |  |  |  | <0.001 |
| <1 | 32 (1.9) | 5 (1.2) | 27 (2.1) |  |
| 1-4 | 109 (6.4) | 11 (2.7) | 98 (7.6) |  |
| 5-9 | 238 (13.9) | 42 (10.1) | 196 (15.2) |  |
| 10-19 | 684 (40.0) | 214 (51.6) | 470 (36.3) |  |
| 20-29 | 225 (13.2) | 38 (9.2) | 187 (14.5) |  |
| 30-39 | 118 (6.9) | 27 (6.5) | 91 (7.0) |  |
| 40-49 | 93 (5.4) | 20 (4.8) | 73 (5.6) |  |
| 50+ | 209 (12.2) | 58 (14.0) | 151 (11.7) |  |
| **Female sex** | 802 (47.0) | 185 (44.6) | 617 (47.7) | 0.290 |
| **DENV immune status** |  |  |  | <0.001 |
| Post-primary | 577 (33.8) | 180 (43.4) | 397 (30.7) |  |
| Primary | 153 (9.0) | 30 (7.2) | 123 (9.5) |  |
| Not tested | 978 (57.3) | 205 (49.4) | 773 (59.8) |  |
| **DENV Serotype** |  |  |  | <0.001 |
| 1 | 905 (53.0) | 226 (54.5) | 679 (52.5) |  |
| 2 | 102 (6.0) | 15 (3.6) | 87 (6.7) |  |
| 3 | 149 (8.7) | 18 (4.3) | 131 (10.1) |  |
| 4 | 50 (2.9) | 19 (4.6) | 31 (2.4) |  |
| Unknown | 502 (29.4) | 137 (33.0) | 365 (28.2) |  |
| **Comorbidities** |  |  |  |  |
| Chronic pulmonary disease or asthma | 347 (20.3) | 76 (18.3) | 271 (21.0) | 0.273 |
| Cancer | 27 (1.6) | 6 (1.4) | 21 (1.6) | 0.978 |
| Chronic kidney disease | 12 (0.7) | 5 (1.2) | 7 (0.5) | 0.285 |
| Coronary heart disease | 52 (3.0) | 16 (3.9) | 36 (2.8) | 0.347 |
| Diabetes | 108 (6.3) | 29 (7.0) | 79 (6.1) | 0.601 |
| High cholesterol | 84 (4.9) | 24 (5.8) | 60 (4.6) | 0.420 |
| Hypertension | 156 (9.1) | 41 (9.9) | 115 (8.9) | 0.611 |
| Arthritis | 23 (1.3) | 5 (1.2) | 18 (1.4) | 0.965 |
| Thyroid disease | 74 (4.3) | 16 (3.9) | 58 (4.5) | 0.682 |
| Obesity | 214 (12.5) | 43 (10.4) | 171 (13.2) | 0.148 |
| Gastritis | 28 (1.6) | 9 (2.2) | 19 (1.5) | 0.451 |
| **Warning signs** |  |  |  |  |
| Persistent vomiting | 420 (24.6) | 156 (37.6) | 264 (20.4) | <0.001 |
| Abdominal pain | 995 (58.3) | 328 (79.0) | 667 (51.6) | <0.001 |
| Restlessness | 797 (46.7) | 268 (64.6) | 529 (40.9) | <0.001 |
| Mucosal bleeding | 274 (16.0) | 95 (22.9) | 179 (13.8) | <0.001 |
| Hemoconcentration | 129 (7.6) | 86 (20.7) | 43 (3.3) | <0.001 |
| Hepatomegaly | 47 (2.8) | 19 (4.6) | 28 (2.2) | 0.015 |
| **Other clinical signs/symptoms** |  |  |  |  |
| Fever | 1695 (99.2) | 412 (99.3) | 1283 (99.2) | 1.000 |
| Conjunctivitis | 147 (8.6) | 26 (6.3) | 121 (9.4) | 0.064 |
| Chills | 1362 (79.7) | 369 (88.9) | 993 (76.8) | <0.001 |
| Nausea | 1175 (68.8) | 333 (80.2) | 842 (65.1) | <0.001 |
| No appetite | 1330 (77.9) | 363 (87.5) | 967 (74.8) | <0.001 |
| Rash | 971 (56.9) | 286 (68.9) | 685 (53.0) | <0.001 |
| Yellow skin | 69 (4.0) | 36 (8.7) | 33 (2.6) | <0.001 |
| Itchy skin | 627 (36.7) | 190 (45.8) | 437 (33.8) | <0.001 |
| Bruise | 103 (6.0) | 35 (8.4) | 68 (5.3) | 0.025 |
| Headache | 1432 (83.8) | 372 (89.6) | 1060 (82.0) | <0.001 |
| Eye pain | 1026 (60.1) | 288 (69.4) | 738 (57.1) | <0.001 |
| Myalgia | 1276 (74.7) | 362 (87.2) | 914 (70.7) | <0.001 |
| Arthralgia | 1062 (62.2) | 301 (72.5) | 761 (58.9) | <0.001 |
| Back pain | 912 (53.4) | 261 (62.9) | 651 (50.3) | <0.001 |
| Calf pain | 627 (36.7) | 180 (43.4) | 447 (34.6) | <0.001 |
| Arthritis | 251 (14.7) | 84 (20.2) | 167 (12.9) | <0.001 |
| Nasal discharge | 545 (31.9) | 149 (35.9) | 396 (30.6) | 0.052 |
| Sore throat | 615 (36.0) | 168 (40.5) | 447 (34.6) | 0.034 |
| Cough | 726 (42.5) | 189 (45.5) | 537 (41.5) | 0.167 |
| Diarrhea | 736 (43.1) | 228 (54.9) | 508 (39.3) | <0.001 |
| Seizure | 30 (1.8) | 30 (7.2) | 0 (0.0) | <0.001 |
| Pale skin | 783 (45.8) | 266 (64.1) | 517 (40.0) | <0.001 |
| Blue lips | 72 (4.2) | 35 (8.4) | 37 (2.9) | <0.001 |
| **Laboratory** |  |  |  |  |
| Leukopenia | 1012 (59.3) | 320 (77.1) | 692 (53.5) | <0.001 |

*p*-values were calculated using either the chi-square test or Fisher's exact test, as appropriate, based on the sample sizes in each category.

### Performance of warning signs for predicting severe dengue

Among warning signs, abdominal pain and restlessness had the highest sensitivities for predicting severe dengue at 79.0% and 64.6%, respectively, but the lowest specificities of 48.4% and 59.1% (Table 2). In contrast, hepatomegaly and hemoconcentration demonstrated the highest specificities at 97.8% and 96.7%, respectively, but were less sensitive at 4.6% and 20.7%, respectively. The presence of any warning sign yielded the highest sensitivity (92.8%) but a low specificity (29.2%), with an AUC-ROC of 61.1%. Combining three or more warning signs increased the specificity to 65.1% while maintaining moderate sensitivity (87.2%), resulting in the highest AUC-ROC (71.3%) among the combinations tested.

**Table 2.** Performance of warning signs for predicting severe dengue (n=1708), Sentinel Enhanced Dengue Surveillance System, Puerto Rico, 2012–2024.

| Warning sign | True Positive<br>n (%) | True Negative<br>n (%) | False Positive<br>n (%) | False Negative<br>n (%) | Sensitivity<br>% (95% CI) | Specificity<br>% (95% CI) | Positive Predictive Value<br>% (95% CI) | Negative Predictive Value<br>% (95% CI) | AUC-ROC<br>% (95% CI) |
| --- | --- | --- | --- | --- | --- | --- | --- | --- | --- |
| Persistent vomiting | 156 (9.1) | 1029 (60.2) | 264 (15.5) | 259 (15.2) | 37.6 (32.9, 42.4) | 79.6 (77.3, 81.7) | 37.1 (32.5, 42.0) | 79.9 (77.6, 82.0) | 58.5 (56.0, 61.1) |
| Abdominal pain | 328 (19.2) | 626 (36.7) | 667 (39.1) | 87 (5.1) | 79.0 (74.8, 82.9) | 48.4 (45.7, 51.2) | 33.0 (30.0, 36.0) | 87.8 (85.2, 90.1) | 60.4 (58.5, 62.3) |
| Restlessness | 268 (15.7) | 764 (44.7) | 529 (31.0) | 147 (8.6) | 64.6 (59.8, 69.2) | 59.1 (56.4, 61.8) | 33.6 (30.3, 37.0) | 83.9 (81.3, 86.2) | 58.7 (56.7, 60.8) |
| Mucosal bleeding | 95 (5.6) | 1114 (65.2) | 179 (10.5) | 320 (18.7) | 22.9 (18.9, 27.2) | 86.2 (84.2, 88.0) | 34.7 (29.0, 40.6) | 77.7 (75.4, 79.8) | 56.2 (53.2, 59.2) |
| Hemoconcentration | 86 (5.0) | 1250 (73.2) | 43 (2.5) | 329 (19.3) | 20.7 (16.9, 24.9) | 96.7 (95.5, 97.6) | 66.7 (57.8, 74.7) | 79.2 (77.1, 81.1) | 72.9 (68.7, 77.1) |
| Hepatomegaly | 19 (1.1) | 1265 (74.1) | 28 (1.6) | 396 (23.2) | 4.6 (2.8, 7.1) | 97.8 (96.9, 98.6) | 40.4 (26.4, 55.7) | 76.2 (74.0, 78.2) | 58.3 (51.1, 65.5) |
| Any warning sign | 388 (22.7) | 347 (20.3) | 946 (55.4) | 27 (1.6) | 93.5 (90.7, 95.7) | 26.8 (24.4, 29.3) | 29.1 (26.7, 31.6) | 92.8 (89.7, 95.2) | 60.9 (59.1, 62.7) |
| Only one warning sign | 78 (9.0) | 347 (40.1) | 413 (47.7) | 27 (3.1) | 74.3 (64.8, 82.3) | 45.7 (42.1, 49.3) | 15.9 (12.8, 19.4) | 92.8 (89.7, 95.2) | 54.3 (52.2, 56.4) |
| Only two warning signs | 126 (14.9) | 347 (41.0) | 347 (41.0) | 27 (3.2) | 82.4 (75.4, 88.0) | 50.0 (46.2, 53.8) | 26.6 (22.7, 30.9) | 92.8 (89.7, 95.2) | 59.7 (57.3, 62.1) |
| Three or more warning signs | 184 (24.7) | 347 (46.6) | 186 (25.0) | 27 (3.6) | 87.2 (81.9, 91.4) | 65.1 (60.9, 69.2) | 49.7 (44.5, 54.9) | 92.8 (89.7, 95.2) | 71.3 (68.4, 74.1) |

**AUC-ROC:** area under receiver operating characteristic curve.

Performance of warning signs for predicting severe dengue demonstrated some variability across serotypes and immune status, though the interpretation is constrained by limited sample sizes and overlapping confidence intervals (Tables S2-S3).

### Performance evaluation of machine learning models

The ensemble model demonstrated a strong correlation between predictions from the CatBoost, XGBoost, LightGBM, and AdaBoost models, with Pearson correlation coefficients of 0.91, 0.89, 0.89, and 0.84, respectively, indicating that these gradient boosting models had substantial influence on the ensemble’s predictions (Figure 2). This high correlation suggests that the models may be selecting and emphasizing similar variables in their predictive processes. In contrast, weaker correlations were observed between the ensemble model and simpler models like KNN (0.48), Naïve Bayes (0.48), and DT (0.51), indicating different prediction patterns and potential differences in variable selection. Additionally, high inter-model correlations among gradient boosting models, particularly between LightGBM and XGBoost (0.98), further support the idea that these models capture similar patterns in the data and rely on comparable sets of variables.

**Figure 2.**
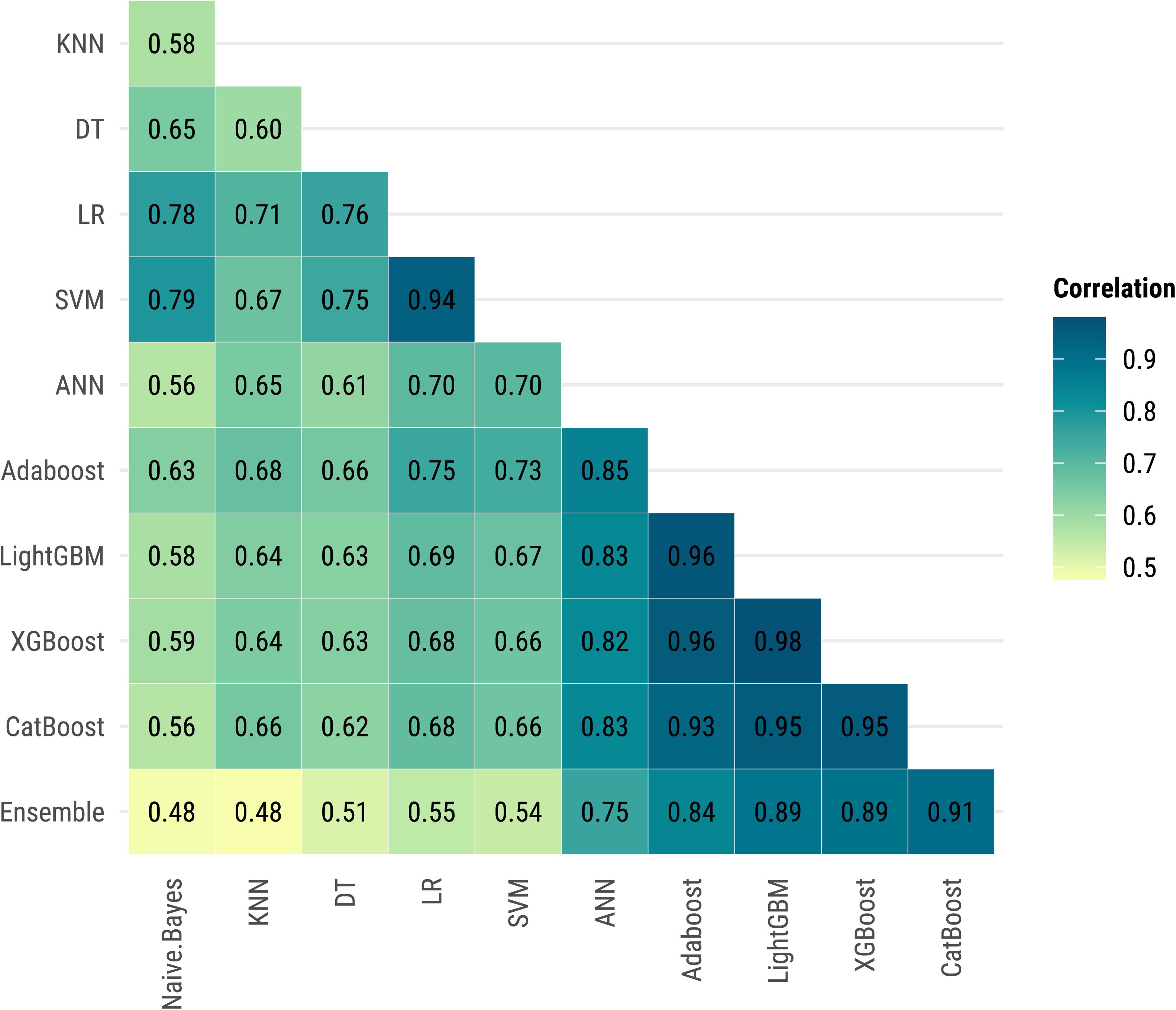
Pearson’s correlation of predictions between machine learning models, Sentinel Enhanced Dengue Surveillance System, Puerto Rico, 2012–2024. Pearson correlation coefficients measure the linear agreement between the predictions of different machine learning models. Higher values indicate similar prediction patterns across models, suggesting that models are identifying similar cases as severe dengue. Darker colors represent higher correlations.

AUC values for the 40-variable feature set across various ML models demonstrated varying levels of predictive performance. Gradient boosting algorithms achieved the highest AUC values of 97.1% for CatBoost, 95.5% for XGBoost, and 94.5% for LightGBM, indicating strong discriminatory power (Figure 3). ANN showed moderate performance (AUC = 88.4%), whereas LR and SVM had lower discrimination (AUC = 79.4% and 78.9%, respectively). KNN, Naïve Bayes, and DT had the lowest AUC values of 74.1%, 75.9%, and 76.2%, respectively, indicating limited predictive ability. The ensemble meta-model provided a slight improvement over CatBoost with an AUC of 97.7%.

**Figure 3.**
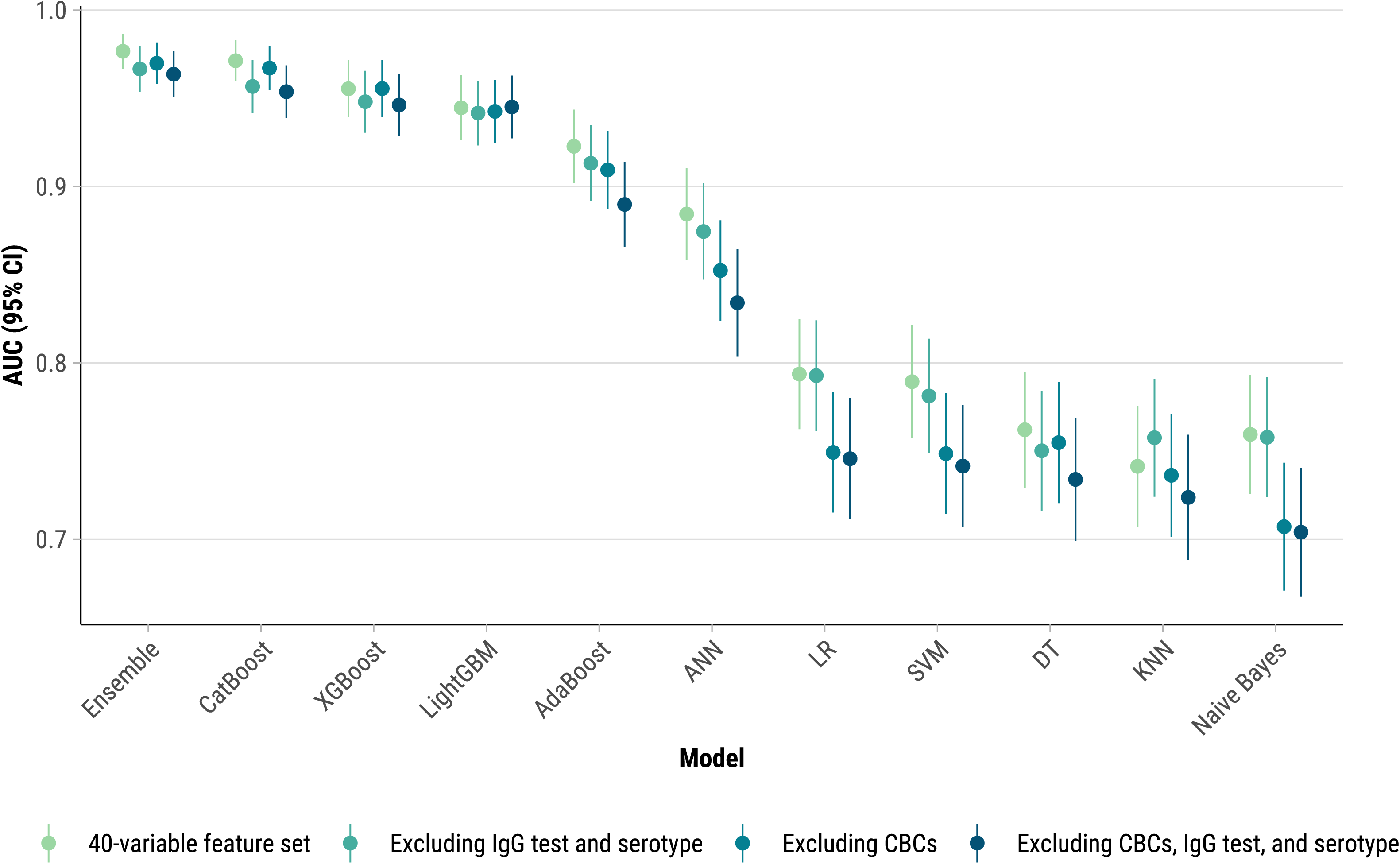
Forest plot of AUC values for Decision Trees (DT), K-Nearest Neighbors (KNN), Naïve Bayes, Support Vector Machines (SVM), Artificial Neural Networks (ANN), Adaptive Boosting (AdaBoost), Categorical Boosting (CatBoost), Light Gradient Boosting Machine (LightGBM), eXtreme Gradient Boosting (XGBoost), and ensemble models for a 40-variable feature set and subsets excluding CBCs, IgG, and serotype results, Sentinel Enhanced Dengue Surveillance System, Puerto Rico, 2012–2024. DeLong method was used to obtain the 95% confidence intervals for the AUC-ROC. CBC = complete blood count, IgG = immunoglobulin G, AUC-ROC = area under the receiver operating characteristic curve.

Exclusion of immune status and serotype data minimally affected model performance across all ML algorithms (Figure 3). Conversely, removing leukopenia and hemoconcentration significantly reduced predictive power for Naïve Bayes, LR, SVM, and ANN (AUC decreased by 3.2% ∼ 5.2%). CatBoost, XGBoost, LightGBM, and the ensemble model consistently maintained high performance, showing minimal to no change in AUC, regardless of the inclusion or exclusion of leukopenia and hemoconcentration. Excluding CHIKV-positive cases in the sensitivity analysis resulted in minimal changes to the AUC-ROC scores for CatBoost, confirming that the model’s predictive performance for severe dengue remains robust even in the presence of co-circulating arboviruses (Table S4).

The ensemble model with 40 variables achieved the highest overall AUC of 97.7% with corresponding sensitivity and specificity of 95.6% and 93.3%, respectively (Table S5). The F1 score was 94.5% and Kappa was 88.9%, indicating a high level of agreement and balanced performance between precision and recall in the model’s classification of severe dengue cases.

### Feature importance

For the 40-variable feature set, SHAP values identified hemoconcentration, days post symptom onset, and leukopenia as most important features for CatBoost, XGBoost, and LightGBM (Figure 4, Figures S1-S2). Similarly, LR highlighted these variables as having the highest adjusted odds ratios for severe dengue (hemoconcentration: aOR 7.02; leukopenia: aOR 2.24; days post onset 4–6 days: aOR 1.96) (Table S6). Additionally, these models highlighted pale skin, age group, and the clinical warning signs of restlessness, abdominal pain, and persistent vomiting as key predictors of severe dengue progression. Hemoconcentration also stood out as a top feature for Naïve Bayes and SVM (Figure S3-S4). AdaBoost, which focuses on correcting errors from previous classifiers, assigned greater importance to chronic conditions such as high cholesterol, chronic arthritis, and hypertension. Although hepatomegaly is a recognized warning sign, it had a lower importance score in our analysis, suggesting it may play a more limited role in predicting severe dengue in this context.

**Figure 4.**
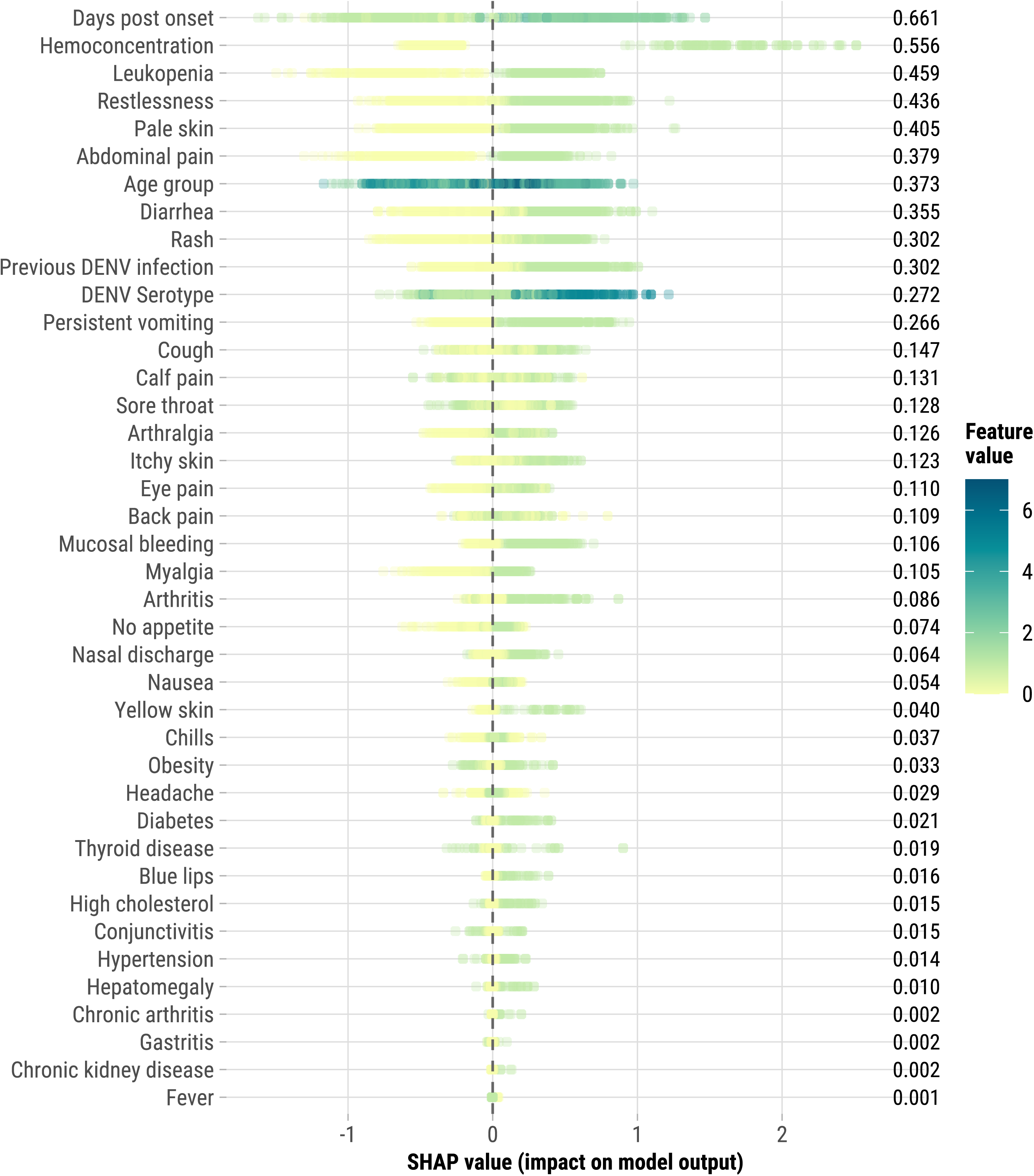
SHapley Additive exPlanations (SHAP) values for the 40 Features in CatBoost, Sentinel Enhanced Dengue Surveillance System, Puerto Rico, 2012–2024. SHAP values measure each feature’s contribution to the prediction of severe dengue in the CatBoost model. Positive SHAP values indicate a higher likelihood of severe dengue, while negative values suggest a lower likelihood (or protective effect). Each dot represents a single case, with its horizontal position showing the SHAP value, reflecting the strength and direction of the feature’s impact. The color of the dots indicates the actual feature value for each case. For most features, values are binary (0 or 1), representing presence or absence (e.g., rash or no rash). For age group, the scale ranges from 0 to 7, with 0 indicating the youngest age group (<1 year) and 7 indicating the oldest age group (≥50 years). An example interpretation: if’ ‘persistent vomiting’ has a positive SHAP value and the dot is green (value = 1), it indicates that the presence of persistent vomiting strongly increases the likelihood of severe dengue for that case. The mean SHAP values shown on the right represent the average absolute impact of each feature across all cases, indicating the overall importance of that feature in the model’s predictions.

### Post-hoc variable reduction analysis

To explore a more streamlined predictive model, we conducted a post-hoc variable reduction analysis using the ML model with the highest AUC, CatBoost. Starting with the top feature identified by SHAP values (days post onset), we sequentially added variables, assessing AUC-ROC at each step. The AUC improved consistently with each additional variable, though the gains diminished over time. By including just 20 variables—compared to the original 40-variable set—the model achieved an AUC of 96.5% (Figure 5). The optimal reduced feature set included days post onset, hemoconcentration, leukopenia, restlessness, pale skin, abdominal pain, age group, diarrhea, rash, persistent vomiting, cough, calf pain, sore throat, arthralgia, itchy skin, eye pain, back pain, mucosal bleeding, myalgia, and arthritis. This reduced model offers a more practical and interpretable approach while maintaining high predictive accuracy, making it feasible for use in clinical settings, especially where diagnostic resources are limited.

**Figure 5.**
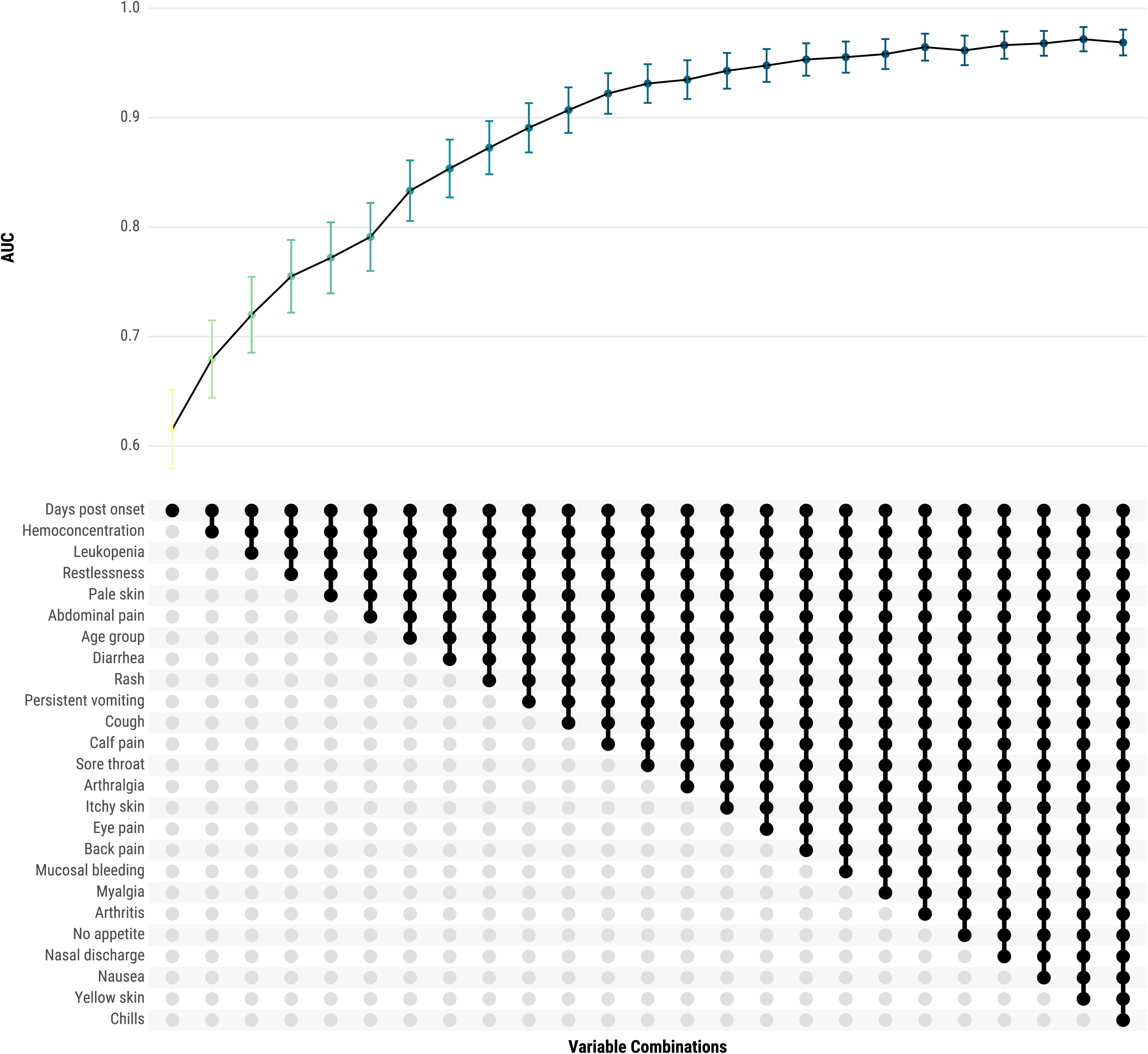
Iterative improvement in area under the curve (AUC) with additional variables in CatBoost model for severe dengue prediction, Sentinel Enhanced Dengue Surveillance System, Puerto Rico, 2012– 2024. This figure shows the change in AUC as top-performing variables are sequentially added to the CatBoost model. Starting with the highest-impact feature, “Days post onset,” each subsequent model includes one additional variable in the order of their mean SHAP values. The combinations of variables and their AUC, along with 95% confidence intervals, are shown to demonstrate the predictive gain with each added variable.

## Discussion

Our study underscores the potential of ML models, particularly gradient boosting algorithms, to outperform traditional warning signs in predicting severe dengue. This improved predictive ability could transform clinical decision-making, enabling earlier and more accurate identification of high-risk patients, thereby improving outcomes in dengue-endemic regions like Puerto Rico.

Hemoconcentration, days post symptom onset, and leukopenia emerged as the most important features across multiple ML models, aligning with their known relevance in dengue prognosis [47–49]. Hemoconcentration, which reflects plasma leakage through an increased red blood cell concentration, was consistently highlighted as a top predictor by CatBoost, XGBoost, LightGBM, and several other algorithms. Days post symptom onset is a crucial temporal marker, likely capturing the dynamic nature of disease as cases often progress to the critical phase of dengue (when severe disease occurs) 3–7 days after symptom onset. Leukopenia, or low white blood cell count, often reflects the body’s response to viral infections, including dengue. In addition, pale skin, age group, and clinical warning signs such as restlessness, abdominal pain, and persistent vomiting were identified as key predictors of severe dengue progression. The variability in feature importance across models emphasizes the complexity of severe dengue prediction, highlighting the need for tailored approaches that account for both individual patient characteristics and disease progression.

Our analysis highlights the strong predictive performance of gradient boosting algorithms— CatBoost, XGBoost, and LightGBM—with AUC values above 94%, reflecting their ability to capture complex, non-linear patterns in clinical data [35, 50–52]. Despite this high predictive accuracy, the interpretability of these models remains a limitation in clinical settings, where transparency in decision-making is critical for trust and practical use [53–55]. Compared to simpler models like LR, which offers straightforward interpretations of how each variable influences severe dengue risk, gradient boosting algorithms can be challenging to interpret. This trade-off between high performance and interpretability suggests that ML models may be most useful as supplementary tools for alerting clinicians to high-risk cases, rather than as standalone decision aids.

In contrast, simpler models like Naïve Bayes, Decision Trees, and KNN showed weaker correlations and lower AUC values, reflecting their limitations in capturing data complexity. The ensemble model, achieving the highest AUC of 97.7%, demonstrates the added value of combining multiple ML algorithms to enhance predictive accuracy, with high sensitivity and specificity, making it particularly useful in resource-limited settings [56, 57]. Additionally, the exclusion of immune status and serotype data had minimal impact on model performance, indicating these variables are not essential for accurate prediction in this context. The high NPV across models suggests that ML tools can still be valuable for identifying low-risk patients who may not require intensive monitoring. In these cases, the ML model’s recommendation could serve as an early discharge or outpatient management decision-support mechanism, further optimizing healthcare resource allocation.

The post-hoc variable reduction analysis demonstrated that a streamlined set of 20 variables achieved strong predictive accuracy (AUC of 96.5%), close to the full 40-variable model. This reduced set offers a balance between interpretability and performance, making it more practical for clinical application, particularly in settings with limited diagnostic resources. By focusing on essential predictors—such as hemoconcentration, days post onset, leukopenia, and key symptoms—this approach prioritizes feasibility and interpretability, even if it means a modest sacrifice in predictive power. For added clinical utility, LR could complement ML approaches by further refining and validating the reduced variable set with interpretable odds ratios, enabling clinicians to apply these findings more confidently in practice [58].

Traditional warning signs showed both strengths and limitations. Abdominal pain and restlessness were the most sensitive indicators, consistent with other studies [6, 9], yet their low specificities limit their utility. Conversely, markers like hepatomegaly and hemoconcentration had high specificity but low sensitivity. Combining multiple warning signs improved specificity while maintaining high sensitivity, yielding the highest AUC among tested combinations. In contrast, gradient boosting ML algorithms offered a more balanced approach with high sensitivity and specificity, crucial for accurate risk stratification in clinical settings. Our findings align with a recent study where an 8-gene XGBoost model outperformed clinical warning signs, significantly improving negative predictive power and demonstrating strong generalizability across patient cohorts [54]. Although our models focus on demographic and clinical features, the integration of gene expression data presents an intriguing avenue for future exploration [54, 55].

This study has several limitations. First, these ML models need to be re-fitted to different variables and populations to ensure accuracy across various settings. Second, the relatively small dataset increases the risk of overfitting, potentially affecting the models’ robustness and generalizability. Third, the prevalence of DENV-1 cases from the 2012-2013 outbreak may limit the models’ applicability to other periods, regions, populations, age groups, or serotypes. Fourth, the models were developed using data from the SEDSS, where inclusion criteria required febrile illness, potentially limiting generalizability to broader populations. Fifth, due to limited sample sizes for serotypes and immune statuses, we could not conduct ML analyses for these subgroups; future studies with larger datasets are needed to validate findings. Sixth, there is a potential limitation related to the inclusion of false negatives—SEDSS cases who may have later presented to a non-SEDSS facility with severe disease—although this is considered unlikely given typical healthcare-seeking behaviors. Seventh, the SEDSS data’s robustness may not accurately reflect real-world conditions, where datasets are often sparse, contain free-text fields, or have incomplete information, potentially affecting model performance. Eighth, dengue and severe dengue are often underdiagnosed and underreported, which could impact model results; however, our findings likely represent a conservative estimate when accounting for underreporting. Finally, implementing ML models in clinical practice may require computational resources and infrastructure not available in all settings, particularly in low-resource environments where dengue is endemic.

## Conclusions

Although traditional warning signs are essential in clinical practice, their low specificity often leads to high hospitalization rates, potentially overwhelming healthcare systems. Our findings suggest that ML models, particularly gradient boosting algorithms, offer a more effective approach by integrating multiple variables and capturing complex interactions, thereby improving specificity while maintaining sensitivity. Implementing these models in clinical decision-making could help identify patients at highest risk for progression to severe dengue, reducing unnecessary hospitalizations and easing healthcare burdens. Although resource constraints may limit direct ML implementation in some settings, platforms like SEDSS can still leverage ML techniques to identify key predictors of severe disease. This approach can optimize patient care by prioritizing the most critical predictors, even in low-resource environments where advanced ML algorithms may not be feasible.

## Abbreviations

AdaBoost: Adaptive Boosting
AFI: acute febrile illness
ANN: Artificial Neural Networks
AUC-ROC: area under the receiver operating characteristic curve
CatBoost: Categorical Boosting
CBC: complete blood count
CHIKV: chikungunya virus
CSPF: convalescent sample processing form
CEMI: Centro de Emergencia y Medicina Integrada
CIF: case investigation form
CMESL: Centro Médico Episcopal San Lucas
DT: Decision Trees
ELISA: enzyme-linked immunosorbent assay
HESL: Hospital Episcopal San Lucas
KNN: K-Nearest Neighbors
LightGBM: Light Gradient Boosting Machine
LR: logistic regression
ML: machine learning
NPV: negative predictive value
PPV: positive predictive value
RT-PCR: reverse transcription polymerase chain reaction
SEDSS: Sentinel Enhanced Dengue Surveillance System
SHAP: SHapley Additive exPlanations
SVM: Support Vector Machines
XGBoost: eXtreme Gradient Boosting
WHO: World Health Organization

## Declarations

### Consent for publication

Not applicable.

### Availability of data and materials

Data cannot be shared publicly because data cannot be deidentified at the granular level of analyses performed. Data are available from the CDC management team (contact) for researchers who meet the criteria for access to confidential data.

### Authors’ contributions

Conception and design of the study: ZJM, MBT, and JMW. Acquisition of data: VR. Analysis and interpretation of data: ZJM. Drafting the article: ZJM. Revising the article critically for important intellectual content: ZJM, DR, MBT, VR, GP, LEA, and JMW. All authors have made substantial contributions to the study. All authors read and approved the final manuscript.

## Acknowledgements

Not applicable.

## Disclaimer

The findings and conclusions in this report are those of the authors and do not necessarily represent the official position of the US Centers for Disease Control and Prevention.

