## Supplement for "Machine learning for predicting severe dengue, Puerto Rico"

**Table S1**. Machine learning models and hyperparameter tuning.

**Table S2**. Performance of warning signs for predicting severe dengue by immune status, Sentinel Enhanced Dengue Surveillance System, Puerto Rico, 2012–2024.

**Table S3**. Performance of warning signs for predicting severe dengue by serotype, Sentinel Enhanced Dengue Surveillance System, Puerto Rico, 2012–2024.

**Table S4**. Performance of CatBoost model for predicting severe dengue on the test set excluding IgM or PCR-positive chikungunya cases, Sentinel Enhanced Dengue Surveillance System, Puerto Rico, 2012–2024.

**Table S5**. Performance of ensemble model for predicting severe dengue on the test set, Sentinel Enhanced Dengue Surveillance System, Puerto Rico, 2012–2024.

**Table S6.** Odds of severe dengue for selected characteristics, Sentinel Enhanced Dengue Surveillance System, Puerto Rico, 2012–2024.

**Figure S1.** Feature importance for each machine learning model for the 40-variable feature set, Sentinel Enhanced Dengue Surveillance System, Puerto Rico, 2012–2024.

**Figure S2.** Feature importance for each machine learning model for a reduced feature set which excludes leukopenia, hemoconcentration, DENV immune status, and DENV serotype, Sentinel Enhanced Dengue Surveillance System, Puerto Rico, 2012–2024.

**Figure S3.** SHapley Additive exPlanations (SHAP) values for the 40 Features in XGBoost, Sentinel Enhanced Dengue Surveillance System, Puerto Rico, 2012–2024.

**Figure S4.** SHapley Additive exPlanations (SHAP) values for the 40 Features in LightGBM, Sentinel Enhanced Dengue Surveillance System, Puerto Rico, 2012–2024.

| **Table S1. Machine learning models and hyperparameter tuning.** To optimize the performance of various machine learning models for predicting severe dengue, a grid search strategy was employed across several algorithms. Specific hyperparameters evaluated for each model are outlined below. We also included the optimal values identified through this process that were included in the final models. | | | |
| --- | --- | --- | --- |
| Model, R package | Parameter | Definition | Included in final model for 40-variable feature set |
| XGBoost,  xgboost [43] | booster type | specifies type of model to use | decision tree |
|  | objective | defines the learning task and the corresponding loss function | binary:logistic |
|  | nrounds | number of boosting iterations | 500 |
|  | max_depth | maximum depth of a tree | 11 |
|  | eta | learning rate | 0.19 |
|  | gamma | minimum loss reduction required to make a further partition on a leaf node of the tree | 0 |
|  | colsample_bytree | subsample ratio of columns when constructing each tree | 0.75 |
|  | min_child_weight | minimum sum of instance weight (hessian) needed in a child | 1 |
| LightGBM,  lightgbm [42] | booster type | specifies type of model to use | decision tree |
|  | objective | defines the learning task | binary |
|  | num_leaves | maximum tree leaves for base learners | 54 |
|  | max_depth | maximum depth of a tree | -1 (no limit) |
|  | learning_rate | shrinks the contribution of each tree by learning_rate | 0.2 |
|  | nrounds | number of boosting iterations | 500 |
|  | min_split_gain | minimum gain to make a split | 0 |
|  | feature_fraction | randomly select a subset of features for each tree during training | 0.8 |
|  | bagging_fraction | randomly sample fraction of data used for each training iteration | 0.9 |
|  | bagging_freq | perform bagging at every k iteration | 8 |
| CatBoost, catboost [41] | loss_function | specifies the loss function | Logloss |
|  | iterations | number of boosting iterations | 500 |
|  | depth | depth of the tree | 12 |
|  | learning_rate | learning rate | 0.1 |
|  | l2_leaf_reg | L2 regularization term on weights | 1 |
|  | border_count | number of splits for numerical features | auto |
| AdaBoost, ada [40] | type | type of boosting algorithm | discrete boosting |
|  | nu | shrinkage parameter for boosting | 0.1 |
|  | iter | number of boosting iterations | 500 |
|  | loss | loss function used to evaluate the model’s performance | exponential |
| SVM, e1071 [37] | kernel type | specifies kernel function used to map data into a higher-dimensional space | linear |
|  | Gamma | Kernel coefficient | 1/n_features = 0.02 |
|  | Cost | controls trade-off between maximizing the margin and minimizing classification errors | 1 |
|  | probability | enables probability predictions | TRUE |
| KNN, caret [26] | k | number of neighbors to consider | 7 |
|  | tunelength | amount of granularity in the tuning parameter grid | 10 |
| Naïve Bayes, e1071 [37] | laplace | Laplace smoothing parameter | 0 |
| DT, rpart [36] | cp | complexity parameter | 0.001 |
|  | Minsplit | minimum number of observations that must exist in a node in order for a split to be considered | 20 |
|  | minbucket | minimum number of observations in any terminal node | 10 |
|  | Maxdepth | maximum depth of any node of the tree | 5 |
|  | method | specifies type of problem | class |
| ANN, nnet [38] | size | number of units in the hidden layer | 15 |
|  | decay | weight decay for regularization to avoid overfitting | 0.2 |
|  | rang | initial range of weights | 0.7 |
|  | maxit | maximum number of iterations | 500 |
|  | entropy | maximum conditional likelihood | least-squares error |

| **Table S2.** Performance of warning signs for predicting severe dengue by immune status, Sentinel Enhanced Dengue Surveillance System, Puerto Rico, 2012–2024. | | | | | | | | | |
| --- | --- | --- | --- | --- | --- | --- | --- | --- | --- |
| **Warning sign** | **True Positive**  n (%) | **True Negative**  n (%) | **False Positive**  n (%) | **False Negative**  n (%) | **Sensitivity**  % (95% CI) | **Specificity**  % (95% CI) | **Positive Predictive Value**  % (95% CI) | **Negative Predictive Value**  % (95% CI) | **AUC-ROC**  % (95% CI) |
| *Post-primary infections* |  |  |  |  |  |  |  |  |  |
| Persistent vomiting | 75 (13.0) | 303 (52.5) | 94 (16.3) | 105 (18.2) | 41.7 (34.4, 49.2) | 76.3 (71.8, 80.4) | 44.4 (36.8, 52.2) | 74.3 (69.7, 78.4) | 59.3 (55.0, 63.6) |
| Abdominal pain | 149 (25.8) | 160 (27.7) | 237 (41.1) | 31 (5.4) | 82.8 (76.5, 88.0) | 40.3 (35.4, 45.3) | 38.6 (33.7, 43.7) | 83.8 (77.8, 88.7) | 61.2 (57.6, 64.8) |
| Restlessness | 115 (19.9) | 240 (41.6) | 157 (27.2) | 65 (11.3) | 63.9 (56.4, 70.9) | 60.5 (55.5, 65.3) | 42.3 (36.3, 48.4) | 78.7 (73.7, 83.1) | 60.5 (56.7, 64.2) |
| Mucosal bleeding | 37 (6.4) | 350 (60.7) | 47 (8.1) | 143 (24.8) | 20.6 (14.9, 27.2) | 88.2 (84.6, 91.2) | 44.0 (33.2, 55.3) | 71.0 (66.8, 75.0) | 57.5 (51.8, 63.2) |
| Hemoconcentration | 40 (6.9) | 377 (65.3) | 20 (3.5) | 140 (24.3) | 22.2 (16.4, 29.0) | 95.0 (92.3, 96.9) | 66.7 (53.3, 78.3) | 72.9 (68.9, 76.7) | 69.8 (63.5, 76.1) |
| Hepatomegaly | 8 (1.4) | 394 (68.3) | 3 (0.5) | 172 (29.8) | 4.4 (1.9, 8.6) | 99.2 (97.8, 99.8) | 72.7 (39.0, 94.0) | 69.6 (65.6, 73.4) | 71.2 (57.2, 85.1) |
| Any warning sign | 173 (30.0) | 87 (15.1) | 310 (53.7) | 7 (1.2) | 96.1 (92.2, 98.4) | 21.9 (17.9, 26.3) | 35.8 (31.5, 40.3) | 92.6 (85.3, 97.0) | 64.2 (60.8, 67.6) |
| Only one warning sign | 37 (14.2) | 87 (33.3) | 130 (49.8) | 7 (2.7) | 84.1 (69.9, 93.4) | 40.1 (33.5, 46.9) | 22.2 (16.1, 29.2) | 92.6 (85.3, 97.0) | 57.4 (53.2, 61.5) |
| Only two warning signs | 53 (19.8) | 87 (32.5) | 121 (45.1) | 7 (2.6) | 88.3 (77.4, 95.2) | 41.8 (35.0, 48.8) | 30.5 (23.7, 37.9) | 92.6 (85.3, 97.0) | 61.5 (57.2, 65.9) |
| Three or more warning signs | 83 (35.2) | 87 (36.9) | 59 (25.0) | 7 (3.0) | 92.2 (84.6, 96.8) | 59.6 (51.2, 67.6) | 58.5 (49.9, 66.7) | 92.6 (85.3, 97.0) | 75.5 (70.6, 80.4) |
| *Primary infections* |  |  |  |  |  |  |  |  |  |
| Persistent vomiting | 16 (10.5) | 92 (60.1) | 31 (20.3) | 14 (9.2) | 53.3 (34.3, 71.7) | 74.8 (66.2, 82.2) | 34.0 (20.9, 49.3) | 86.8 (78.8, 92.6) | 60.4 (52.8, 68.0) |
| Abdominal pain | 27 (17.6) | 62 (40.5) | 61 (39.9) | 3 (2.0) | 90.0 (73.5, 97.9) | 50.4 (41.2, 59.5) | 30.7 (21.3, 41.4) | 95.4 (87.1, 99.0) | 63.0 (57.5, 68.5) |
| Restlessness | 16 (10.5) | 76 (49.7) | 47 (30.7) | 14 (9.2) | 53.3 (34.3, 71.7) | 61.8 (52.6, 70.4) | 25.4 (15.3, 37.9) | 84.4 (75.3, 91.2) | 54.9 (48.3, 61.5) |
| Mucosal bleeding | 6 (3.9) | 113 (73.9) | 10 (6.5) | 24 (15.7) | 20.0 (7.7, 38.6) | 91.9 (85.6, 96.0) | 37.5 (15.2, 64.6) | 82.5 (75.1, 88.4) | 60.0 (47.3, 72.7) |
| Hemoconcentration | 4 (2.6) | 118 (77.1) | 5 (3.3) | 26 (17.0) | 13.3 (3.8, 30.7) | 95.9 (90.8, 98.7) | 44.4 (13.7, 78.8) | 81.9 (74.7, 87.9) | 63.2 (45.7, 80.7) |
| Hepatomegaly | 2 (1.3) | 122 (79.7) | 1 (0.7) | 28 (18.3) | 6.7 (0.8, 22.1) | 99.2 (95.6, 100.0) | 66.7 (9.4, 99.2) | 81.3 (74.2, 87.2) | 74.0 (41.2, 100.0) |
| Any warning sign | 29 (19.0) | 38 (24.8) | 85 (55.6) | 1 (0.7) | 96.7 (82.8, 99.9) | 30.9 (22.9, 39.9) | 25.4 (17.7, 34.4) | 97.4 (86.5, 99.9) | 61.4 (56.7, 66.2) |
| Only one warning sign | 6 (7.4) | 38 (46.9) | 36 (44.4) | 1 (1.2) | 85.7 (42.1, 99.6) | 51.4 (39.4, 63.1) | 14.3 (5.4, 28.5) | 97.4 (86.5, 99.9) | 55.9 (49.9, 61.8) |
| Only two warning signs | 12 (14.5) | 38 (45.8) | 32 (38.6) | 1 (1.2) | 92.3 (64.0, 99.8) | 54.3 (41.9, 66.3) | 27.3 (15.0, 42.8) | 97.4 (86.5, 99.9) | 62.4 (55.2, 69.5) |
| Three or more warning signs | 11 (16.4) | 38 (56.7) | 17 (25.4) | 1 (1.5) | 91.7 (61.5, 99.8) | 69.1 (55.2, 80.9) | 39.3 (21.5, 59.4) | 97.4 (86.5, 99.9) | 68.4 (58.8, 77.9) |
| **AUC-ROC**: area under receiver operating characteristic curve. | | | | | | | | | |

| **Table S3.** Performance of warning signs for predicting severe dengue by serotype, Sentinel Enhanced Dengue Surveillance System, Puerto Rico, 2012–2024. | | | | | | | | | |
| --- | --- | --- | --- | --- | --- | --- | --- | --- | --- |
| **Warning sign** | **True Positive**  n (%) | **True Negative**  n (%) | **False Positive**  n (%) | **False Negative**  n (%) | **Sensitivity**  % (95% CI) | **Specificity**  % (95% CI) | **Positive Predictive Value**  % (95% CI) | **Negative Predictive Value**  % (95% CI) | **AUC-ROC**  % (95% CI) |
| *DENV-1* |  |  |  |  |  |  |  |  |  |
| Persistent vomiting | 89 (9.8) | 527 (58.2) | 152 (16.8) | 137 (15.1) | 39.4 (33.0, 46.1) | 77.6 (74.3, 80.7) | 36.9 (30.8, 43.4) | 79.4 (76.1, 82.4) | 58.1 (54.7, 61.6) |
| Abdominal pain | 184 (20.3) | 302 (33.4) | 377 (41.7) | 42 (4.6) | 81.4 (75.7, 86.3) | 44.5 (40.7, 48.3) | 32.8 (28.9, 36.9) | 87.8 (83.9, 91.1) | 60.3 (57.7, 62.9) |
| Restlessness | 142 (15.7) | 413 (45.6) | 266 (29.4) | 84 (9.3) | 62.8 (56.2, 69.1) | 60.8 (57.0, 64.5) | 34.8 (30.2, 39.6) | 83.1 (79.5, 86.3) | 59.0 (56.1, 61.8) |
| Mucosal bleeding | 50 (5.5) | 606 (67.0) | 73 (8.1) | 176 (19.4) | 22.1 (16.9, 28.1) | 89.2 (86.7, 91.5) | 40.7 (31.9, 49.9) | 77.5 (74.4, 80.4) | 59.1 (54.5, 63.7) |
| Hemoconcentration | 52 (5.7) | 655 (72.4) | 24 (2.7) | 174 (19.2) | 23.0 (17.7, 29.1) | 96.5 (94.8, 97.7) | 68.4 (56.7, 78.6) | 79.0 (76.1, 81.7) | 73.7 (68.3, 79.2) |
| Hepatomegaly | 12 (1.3) | 670 (74.0) | 9 (1.0) | 214 (23.6) | 5.3 (2.8, 9.1) | 98.7 (97.5, 99.4) | 57.1 (34.0, 78.2) | 75.8 (72.8, 78.6) | 66.5 (55.5, 77.4) |
| Any warning sign | 214 (23.6) | 180 (19.9) | 499 (55.1) | 12 (1.3) | 94.7 (90.9, 97.2) | 26.5 (23.2, 30.0) | 30.0 (26.7, 33.5) | 93.8 (89.3, 96.7) | 61.9 (59.5, 64.3) |
| Only one warning sign | 48 (10.6) | 180 (39.9) | 211 (46.8) | 12 (2.7) | 80.0 (67.7, 89.2) | 46.0 (41.0, 51.1) | 18.5 (14.0, 23.8) | 93.8 (89.3, 96.7) | 56.1 (53.2, 59.1) |
| Only two warning signs | 65 (14.4) | 180 (40.0) | 193 (42.9) | 12 (2.7) | 84.4 (74.4, 91.7) | 48.3 (43.1, 53.5) | 25.2 (20.0, 31.0) | 93.8 (89.3, 96.7) | 59.5 (56.3, 62.6) |
| Three or more warning signs | 101 (26.0) | 180 (46.4) | 95 (24.5) | 12 (3.1) | 89.4 (82.2, 94.4) | 65.5 (59.5, 71.1) | 51.5 (44.3, 58.7) | 93.8 (89.3, 96.7) | 72.6 (68.7, 76.5) |
| *DENV-2* |  |  |  |  |  |  |  |  |  |
| Persistent vomiting | 4 (3.9) | 73 (71.6) | 14 (13.7) | 11 (10.8) | 26.7 (7.8, 55.1) | 83.9 (74.5, 90.9) | 22.2 (6.4, 47.6) | 86.9 (77.8, 93.3) | 54.6 (44.0, 65.1) |
| Abdominal pain | 14 (13.7) | 51 (50.0) | 36 (35.3) | 1 (1.0) | 93.3 (68.1, 99.8) | 58.6 (47.6, 69.1) | 28.0 (16.2, 42.5) | 98.1 (89.7, 100.0) | 63.0 (56.5, 69.6) |
| Restlessness | 8 (7.8) | 57 (55.9) | 30 (29.4) | 7 (6.9) | 53.3 (26.6, 78.7) | 65.5 (54.6, 75.4) | 21.1 (9.6, 37.3) | 89.1 (78.8, 95.5) | 55.1 (47.4, 62.7) |
| Mucosal bleeding | 5 (4.9) | 67 (65.7) | 20 (19.6) | 10 (9.8) | 33.3 (11.8, 61.6) | 77.0 (66.8, 85.4) | 20.0 (6.8, 40.7) | 87.0 (77.4, 93.6) | 53.5 (44.7, 62.4) |
| Hemoconcentration | 3 (2.9) | 87 (85.3) | 0 (0) | 12 (11.8) | 20.0 (4.3, 48.1) | 100.0 (95.8, 100.0) | 100.0 (29.2, 100.0) | 87.9 (79.8, 93.6) | 93.9 (90.7, 97.2) |
| Hepatomegaly | 2 (2.0) | 82 (80.4) | 5 (4.9) | 13 (12.7) | 13.3 (1.7, 40.5) | 94.3 (87.1, 98.1) | 28.6 (3.7, 71.0) | 86.3 (77.7, 92.5) | 57.4 (39.0, 75.8) |
| Any warning sign | 15 (14.7) | 27 (26.5) | 60 (58.8) | 0 (0) | 100.0 (78.2, 100.0) | 31.0 (21.5, 41.9) | 20.0 (11.6, 30.8) | 100.0 (87.2, 100.0) | 60.0 (55.4, 64.6) |
| Only one warning sign | 3 (4.6) | 27 (41.5) | 35 (53.8) | 0 (0) | 100.0 (29.2, 100.0) | 43.5 (31.0, 56.7) | 7.9 (1.7, 21.4) | 100.0 (87.2, 100.0) | 53.9 (49.6, 58.3) |
| Only two warning signs | 4 (10.0) | 27 (67.5) | 9 (22.5) | 0 (0) | 100.0 (39.8, 100.0) | 75.0 (57.8, 87.9) | 30.8 (9.1, 61.4) | 100.0 (87.2, 100.0) | 65.4 (52.3, 78.4) |
| Three or more warning signs | 8 (15.7) | 27 (52.9) | 16 (31.4) | 0 (0) | 100.0 (63.1, 100.0) | 62.8 (46.7, 77.0) | 33.3 (15.6, 55.3) | 100.0 (87.2, 100.0) | 66.7 (57.0, 76.3) |
| *DENV-3* |  |  |  |  |  |  |  |  |  |
| Persistent vomiting | 4 (2.7) | 115 (77.2) | 16 (10.7) | 14 (9.4) | 22.2 (6.4, 47.6) | 87.8 (80.9, 92.9) | 20.0 (5.7, 43.7) | 89.1 (82.5, 93.9) | 54.6 (45.2, 64.0) |
| Abdominal pain | 13 (8.7) | 75 (50.3) | 56 (37.6) | 5 (3.4) | 72.2 (46.5, 90.3) | 57.3 (48.3, 65.9) | 18.8 (10.4, 30.1) | 93.8 (86.0, 97.9) | 56.3 (50.9, 61.7) |
| Restlessness | 10 (6.7) | 89 (59.7) | 42 (28.2) | 8 (5.4) | 55.6 (30.8, 78.5) | 67.9 (59.2, 75.8) | 19.2 (9.6, 32.5) | 91.8 (84.4, 96.4) | 55.5 (49.4, 61.6) |
| Mucosal bleeding | 8 (5.4) | 99 (66.4) | 32 (21.5) | 10 (6.7) | 44.4 (21.5, 69.2) | 75.6 (67.3, 82.7) | 20.0 (9.1, 35.6) | 90.8 (83.8, 95.5) | 55.4 (48.6, 62.3) |
| Hemoconcentration | 1 (0.7) | 131 (87.9) | 0 (0) | 17 (11.4) | 5.6 (0.1, 27.3) | 100.0 (97.2, 100.0) | 100.0 (2.5, 100.0) | 88.5 (82.2, 93.2) | -- |
| Hepatomegaly | 1 (0.7) | 124 (83.2) | 7 (4.7) | 17 (11.4) | 5.6 (0.1, 27.3) | 94.7 (89.3, 97.8) | 12.5 (0.3, 52.7) | 87.9 (81.4, 92.8) | 50.2 (37.7, 62.8) |
| Any warning sign | 15 (10.1) | 41 (27.5) | 90 (60.4) | 3 (2.0) | 83.3 (58.6, 96.4) | 31.3 (23.5, 40.0) | 14.3 (8.2, 22.5) | 93.2 (81.3, 98.6) | 53.7 (48.7, 58.8) |
| Only one warning sign | 1 (1.1) | 41 (47.1) | 42 (48.3) | 3 (3.4) | 25.0 (0.6, 80.6) | 49.4 (38.2, 60.6) | 2.3 (0.1, 12.3) | 93.2 (81.3, 98.6) | 47.8 (43.4, 52.2) |
| Only two warning signs | 8 (9.1) | 41 (46.6) | 36 (40.9) | 3 (3.4) | 72.7 (39.0, 94.0) | 53.2 (41.5, 64.7) | 18.2 (8.2, 32.7) | 93.2 (81.3, 98.6) | 55.7 (48.8, 62.6) |
| Three or more warning signs | 6 (9.7) | 41 (66.1) | 12 (19.4) | 3 (4.8) | 66.7 (29.9, 92.5) | 77.4 (63.8, 87.7) | 33.3 (13.3, 59.0) | 93.2 (81.3, 98.6) | 63.3 (51.4, 75.1) |
| *DENV-4* |  |  |  |  |  |  |  |  |  |
| Persistent vomiting | 8 (16.0) | 26 (52.0) | 5 (10.0) | 11 (22.0) | 42.1 (20.3, 66.5) | 83.9 (66.3, 94.5) | 61.5 (31.6, 86.1) | 70.3 (53.0, 84.1) | 65.9 (50.2, 81.6) |
| Abdominal pain | 16 (32.0) | 16 (32.0) | 15 (30.0) | 3 (6.0) | 84.2 (60.4, 96.6) | 51.6 (33.1, 69.8) | 51.6 (33.1, 69.8) | 84.2 (60.4, 96.6) | 67.9 (55.6, 80.2) |
| Restlessness | 17 (34.0) | 18 (36.0) | 13 (26.0) | 2 (4.0) | 89.5 (66.9, 98.7) | 58.1 (39.1, 75.5) | 56.7 (37.4, 74.5) | 90.0 (68.3, 98.8) | 73.3 (62.1, 84.6) |
| Mucosal bleeding | 4 (8.0) | 24 (48.0) | 7 (14.0) | 15 (30.0) | 21.1 (6.1, 45.6) | 77.4 (58.9, 90.4) | 36.4 (10.9, 69.2) | 61.5 (44.6, 76.6) | 49.0 (32.2, 65.7) |
| Hemoconcentration | 2 (4.0) | 29 (58.0) | 2 (4.0) | 17 (34.0) | 10.5 (1.3, 33.1) | 93.5 (78.6, 99.2) | 50.0 (6.8, 93.2) | 63.0 (47.5, 76.8) | 56.5 (27.4, 85.7) |
| Hepatomegaly | 0 (0) | 31 (62.0) | 0 (0) | 19 (38.0) | 0.0 (0.0, 17.6) | 100.0 (88.8, 100.0) | -- | 62.0 (47.2, 75.3) | -- |
| Any warning sign | 19 (38.0) | 11 (22.0) | 20 (40.0) | 0 (0) | 100.0 (82.4, 100.0) | 35.5 (19.2, 54.6) | 48.7 (32.4, 65.2) | 100.0 (71.5, 100.0) | 74.4 (66.4, 82.3) |
| Only one warning sign | 2 (10.0) | 11 (55.0) | 7 (35.0) | 0 (0) | 100.0 (15.8, 100.0) | 61.1 (35.7, 82.7) | 22.2 (2.8, 60.0) | 100.0 (71.5, 100.0) | 61.1 (46.7, 75.5) |
| Only two warning signs | 9 (34.6) | 11 (42.3) | 6 (23.1) | 0 (0) | 100.0 (66.4, 100.0) | 64.7 (38.3, 85.8) | 60.0 (32.3, 83.7) | 100.0 (71.5, 100.0) | 80.0 (67.2, 92.8) |
| Three or more warning signs | 8 (30.8) | 11 (42.3) | 7 (26.9) | 0 (0) | 100.0 (63.1, 100.0) | 61.1 (35.7, 82.7) | 53.3 (26.6, 78.7) | 100.0 (71.5, 100.0) | 76.7 (63.6, 89.7) |
| **AUC-ROC**: area under receiver operating characteristic curve. | | | | | | | | | |

| **Table S4.** Performance of CatBoost model for predicting severe dengue on the test set excluding IgM or PCR-positive chikungunya cases, Sentinel Enhanced Dengue Surveillance System, Puerto Rico, 2012–2024. | | | | | | | | |
| --- | --- | --- | --- | --- | --- | --- | --- | --- |
| **Model** | **Accuracy**  **%** | **Sensitivity**  **%** | **Specificity**  **%** | **PPV**  **%** | **NPV**  **%** | **F1 Score**  **%** | **Kappa**  **%** | **AUC**  **%** |
| 40-variable feature set | 93.3 | 95.4 | 91.3 | 91.4 | 95.3 | 93.4 | 86.7 | 96.6 |
| Excluding leukopenia and hemoconcentration | 91.1 | 91.3 | 90.8 | 90.6 | 91.5 | 91.0 | 82.1 | 95.6 |
| Excluding immune status and serotype results | 91.5 | 91.3 | 91.6 | 91.3 | 91.6 | 91.3 | 82.9 | 96.0 |
| Excluding leukopenia, hemoconcentration, serotype, and immune status | 90.3 | 88.1 | 92.4 | 91.8 | 88.9 | 89.9 | 80.5 | 95.0 |
| **PPV:** positive predictive value; **NPV**: negative predictive value; **AUC**: area under receiver operating characteristic curve. | | | | | | | | |

| **Table S5.** Performance of ensemble model for predicting severe dengue on the test set, Sentinel Enhanced Dengue Surveillance System, Puerto Rico, 2012–2024. | | | | | | | | |
| --- | --- | --- | --- | --- | --- | --- | --- | --- |
| **Model** | **Accuracy**  **%** | **Sensitivity**  **%** | **Specificity**  **%** | **PPV**  **%** | **NPV**  **%** | **F1 Score**  **%** | **Kappa**  **%** | **AUC**  **%** |
| 40-variable feature set | 94.4 | 95.6 | 93.3 | 93.3 | 95.5 | 94.5 | 88.9 | 97.7 |
| Excluding leukopenia and hemoconcentration | 93.9 | 94.8 | 93.0 | 93.2 | 94.7 | 94.0 | 87.9 | 97.0 |
| Excluding immune status and serotype results | 93.0 | 93.5 | 92.5 | 92.6 | 93.5 | 93.1 | 86.1 | 96.7 |
| Excluding leukopenia, hemoconcentration, serotype, and immune status | 92.9 | 93.8 | 92.0 | 92.1 | 93.7 | 93.0 | 85.8 | 96.4 |
| **PPV:** positive predictive value; **NPV**: negative predictive value; **AUC**: area under receiver operating characteristic curve. | | | | | | | | |

| **Table S6.** Odds of severe dengue for selected characteristics, Sentinel Enhanced Dengue Surveillance System, Puerto Rico, 2012–2024 (N = 1708). | | | | |
| --- | --- | --- | --- | --- |
|  | **OR (95% CI)** | **p-value** | **aOR^a^ (95% CI)** | **p-value** |
| **Days post onset (ref: 0)** |  | <0.001 |  | <0.001 |
| 1-3 | 1.66 (1.06, 2.65) |  | 0.96 (0.57, 1.65) |  |
| 4-6 | 4.69 (3.01, 7.51) |  | 1.96 (1.16, 3.39) |  |
| 7+ | 3.70 (2.00, 6.99) |  | 1.08 (0.50, 2.36) |  |
| **Age group (ref: <1)** |  | <0.001 |  | <0.001 |
| 1-4 | 0.79 (0.31, 2.12) |  | 0.62 (0.21, 1.97) |  |
| 5-9 | 1.77 (0.77, 4.44) |  | 0.90 (0.33, 2.66) |  |
| 10-19 | 3.54 (1.60, 8.64) |  | 1.37 (0.51, 3.95) |  |
| 20-29 | 1.66 (0.72, 4.16) |  | 0.45 (0.16, 1.35) |  |
| 30-39 | 1.84 (0.77, 4.80) |  | 0.67 (0.23, 2.12) |  |
| 40-49 | 2.40 (0.99, 6.28) |  | 0.77 (0.26, 2.43) |  |
| 50+ | 2.99 (1.31, 7.47) |  | 1.33 (0.48, 3.92) |  |
| **DENV serotype (ref: 1)** |  | <0.001 |  | 0.008 |
| 2 | 0.60 (0.37, 0.94) |  | 1.43 (0.80, 2.53) |  |
| 3 | 0.46 (0.31, 0.67) |  | 0.86 (0.53, 1.41) |  |
| 4 | 2.32 (1.38, 4.06) |  | 1.70 (0.92, 3.26) |  |
| Unknown | 1.13 (0.92, 1.40) |  | 1.57 (1.17, 2.13) |  |
| **Immune status (ref: primary infection)** |  | <0.001 |  | 0.007 |
| Post-primary infection | 2.02 (1.42, 2.89) |  | 1.29 (0.83, 2.01) |  |
| Not tested | 1.18 (0.84, 1.68) |  | 0.82 (0.52, 1.31) |  |
| **Comorbidities** |  |  |  |  |
| Chronic kidney disease | 3.03 (1.05, 10.86) | 0.040 | 3.53 (0.98, 14.95) | 0.054 |
| Diabetes | 1.21 (0.83, 1.77) | 0.333 | – | – |
| High cholesterol | 1.24 (0.81, 1.91) | 0.328 | – | – |
| Hypertension | 0.99 (0.72, 1.36) | 0.935 | – | – |
| Arthritis | 0.91 (0.38, 2.16) | 0.826 | 2.09 (0.80, 5.39) | 0.131 |
| Thyroid disease | 0.84 (0.53, 1.35) | 0.476 | – | – |
| Obesity | 0.76 (0.57, 1.01) | 0.061 | – | – |
| Gastritis | 1.38 (0.73, 2.70) | 0.324 | – | – |
| **Warning signs** |  |  |  |  |
| Persistent vomiting | 2.42 (1.95, 2.99) | <0.001 | 1.55 (1.19, 2.02) | 0.001 |
| Abdominal pain | 3.34 (2.72, 4.10) | <0.001 | 1.49 (1.15, 1.94) | 0.003 |
| Restlessness | 2.43 (2.02, 2.94) | <0.001 | 1.81 (1.43, 2.30) | <0.001 |
| Mucosal bleeding | 2.06 (1.61, 2.64) | <0.001 | 1.66 (1.23, 2.25) | 0.001 |
| Hemoconcentration | 8.77 (5.93, 13.45) | <0.001 | 7.02 (4.56, 11.20) | <0.001 |
| Hepatomegaly | 1.67 (0.92, 3.13) | 0.095 | – | – |
| **Other clinical signs** |  |  |  |  |
| Fever | 1.17 (0.39, 3.64) | 0.781 | 0.23 (0.07, 0.81) | 0.022 |
| Conjunctivitis | 0.70 (0.50, 0.97) | 0.034 | – | – |
| Chills | 2.32 (1.79, 3.01) | <0.001 | – | – |
| Nausea | 2.30 (1.86, 2.86) | <0.001 | – | – |
| No appetite | 2.54 (1.99, 3.26) | <0.001 | – | – |
| Rash | 1.79 (1.48, 2.17) | <0.001 | 1.49 (1.16, 1.90) | 0.002 |
| Yellow skin | 2.79 (1.76, 4.59) | <0.001 | 1.50 (0.88, 2.65) | 0.141 |
| Itchy skin | 1.52 (1.26, 1.83) | <0.001 | – | – |
| Headache | 2.07 (1.57, 2.74) | <0.001 | – | – |
| Eye pain | 1.70 (1.40, 2.06) | <0.001 | – | – |
| Myalgia | 3.04 (2.37, 3.91) | <0.001 | 1.82 (1.31, 2.53) | <0.001 |
| Arthralgia | 1.84 (1.51, 2.25) | <0.001 | – | – |
| Back pain | 1.68 (1.40, 2.03) | <0.001 | – | – |
| Calf pain | 1.49 (1.24, 1.80) | <0.001 | 0.76 (0.60, 0.98) | 0.034 |
| Arthritis | 1.68 (1.31, 2.17) | <0.001 | – | – |
| Nasal discharge | 1.17 (0.96, 1.42) | 0.120 | 1.22 (0.94, 1.59) | 0.133 |
| Sore throat | 1.18 (0.98, 1.43) | 0.088 | 0.72 (0.56, 0.93) | 0.010 |
| Cough | 1.18 (0.98, 1.42) | 0.087 | – | – |
| Diarrhea | 2.28 (1.89, 2.75) | <0.001 | 1.40 (1.10, 1.77) | 0.005 |
| Pale skin | 2.67 (2.21, 3.23) | <0.001 | 1.55 (1.22, 1.96) | <0.001 |
| Blue lips | 2.57 (1.67, 4.06) | <0.001 | – | – |
| Leukopenia | 3.19 (2.60, 3.93) | <0.001 | 2.24 (1.70, 2.96) | <0.001 |
| **OR**: odds ratio; **aOR**: adjusted odds ratio. **95% CI**: 95% confidence interval  ^a^ Adjusted for all other variables listed in the model. | | | | |

**Figure S1.** SHapley Additive exPlanations (SHAP) values for the 40 Features in XGBoost, Sentinel Enhanced Dengue Surveillance System, Puerto Rico, 2012–2024. SHAP values measure each feature’s contribution to the prediction of severe dengue in the XGBoost model. Positive SHAP values indicate a higher likelihood of severe dengue, while negative values suggest a lower likelihood (or protective effect). Each dot represents a single case, with its horizontal position showing the SHAP value, reflecting the strength and direction of the feature’s impact. The color of the dots indicates the actual feature value for each case. For most features, values are binary (0 or 1), representing presence or absence (e.g., rash or no rash). For age group, the scale ranges from 0 to 7, with 0 indicating the youngest age group (<1 year) and 7 indicating the oldest age group (≥50 years). An example interpretation: if' ‘persistent vomiting’ has a positive SHAP value and the dot is green (value = 1), it indicates that the presence of persistent vomiting strongly increases the likelihood of severe dengue for that case. The mean SHAP values shown on the right represent the average absolute impact of each feature across all cases, indicating the overall importance of that feature in the model’s predictions.

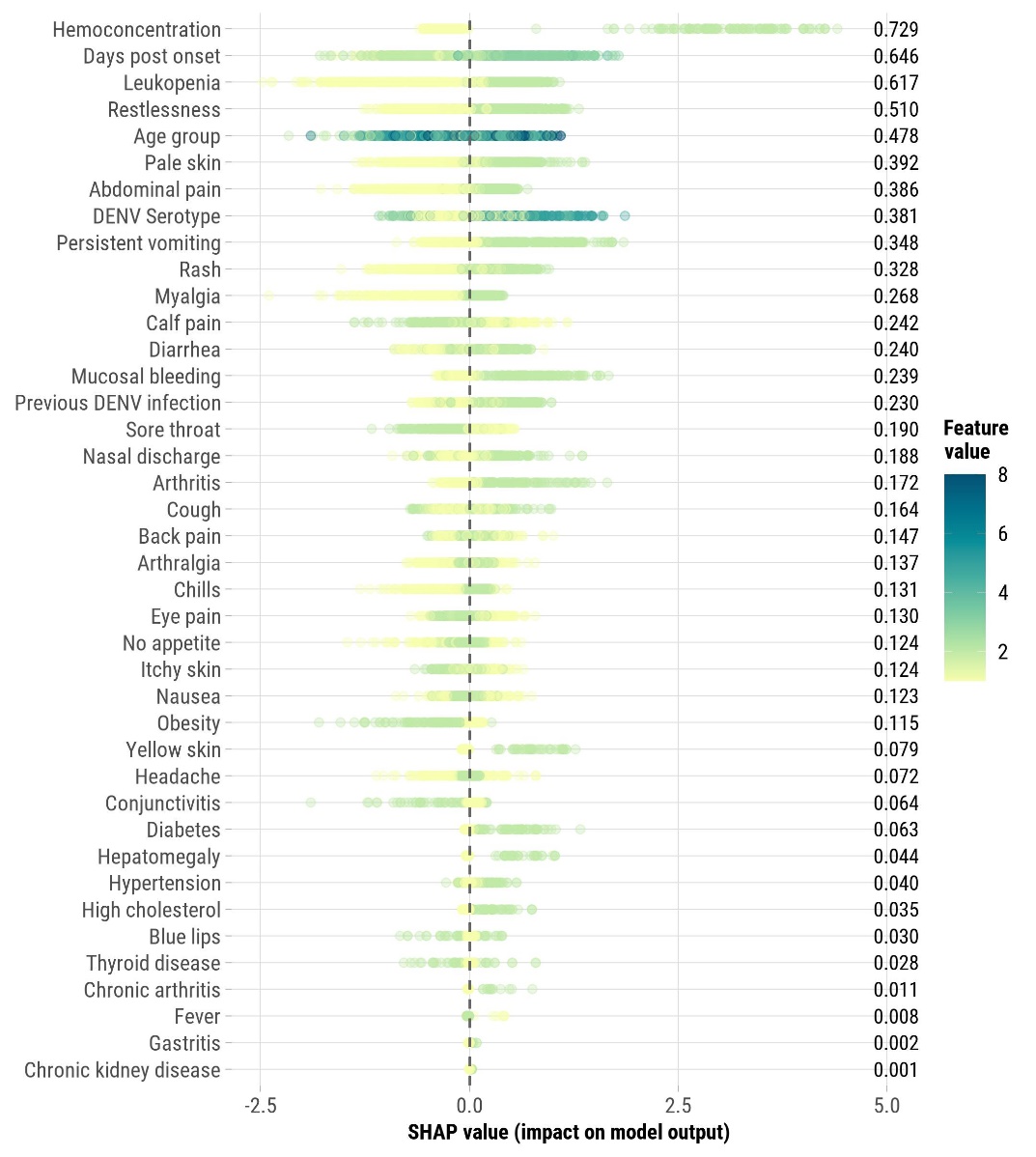

**Figure S2.** SHapley Additive exPlanations (SHAP) values for the 40 Features in LightGBM, Sentinel Enhanced Dengue Surveillance System, Puerto Rico, 2012–2024. SHAP values measure each feature’s contribution to the prediction of severe dengue in the LightGBM model. Positive SHAP values indicate a higher likelihood of severe dengue, while negative values suggest a lower likelihood (or protective effect). Each dot represents a single case, with its horizontal position showing the SHAP value, reflecting the strength and direction of the feature’s impact. The color of the dots indicates the actual feature value for each case. For most features, values are binary (0 or 1), representing presence or absence (e.g., rash or no rash). For age group, the scale ranges from 0 to 7, with 0 indicating the youngest age group (<1 year) and 7 indicating the oldest age group (≥50 years). An example interpretation: if' ‘persistent vomiting’ has a positive SHAP value and the dot is green (value = 1), it indicates that the presence of persistent vomiting strongly increases the likelihood of severe dengue for that case. The mean SHAP values shown on the right represent the average absolute impact of each feature across all cases, indicating the overall importance of that feature in the model’s predictions.

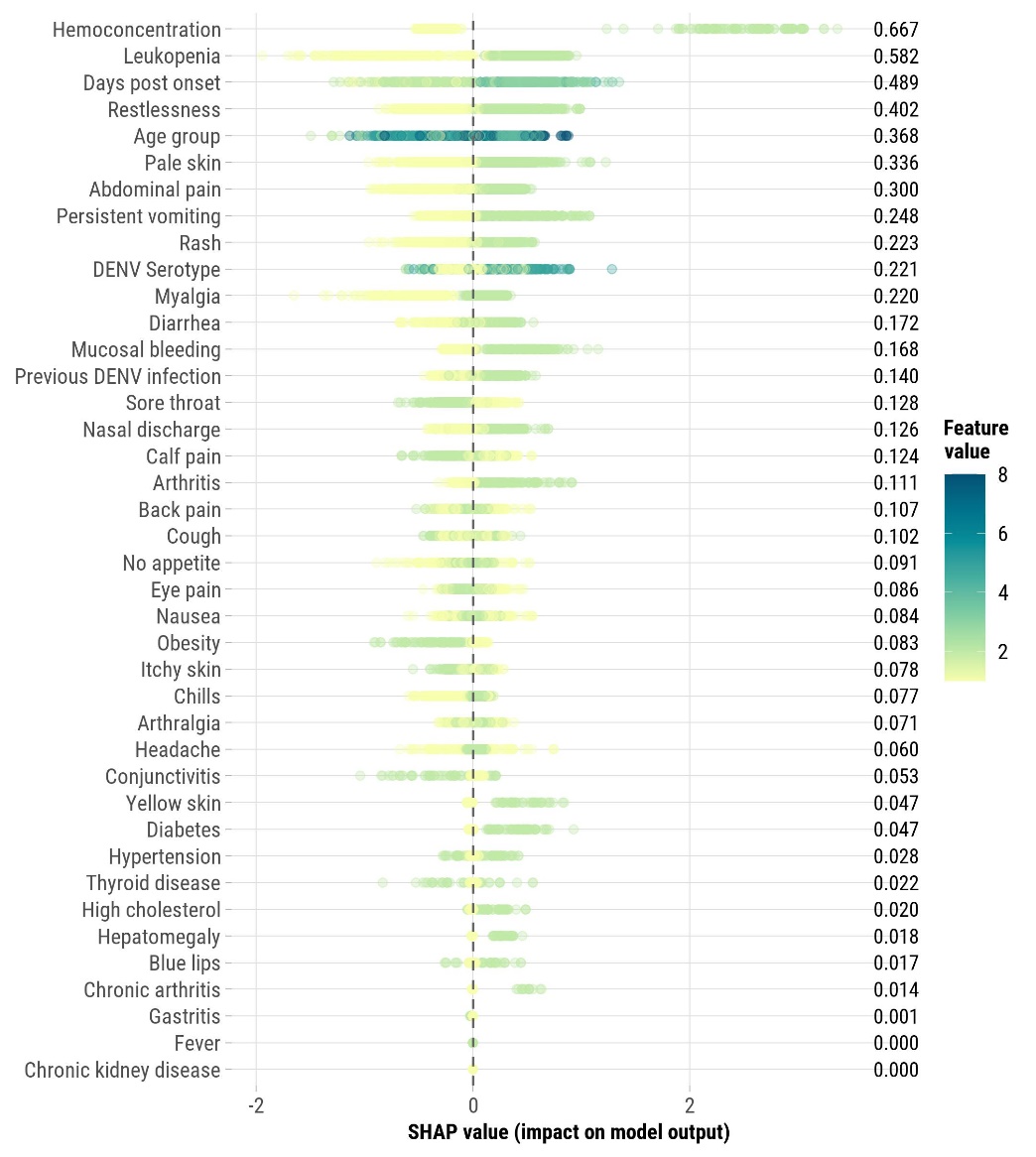

**Figure S3. Feature importance for each machine learning model for the 40-variable feature set, Sentinel Enhanced Dengue Surveillance System, Puerto Rico, 2012–2024.** The feature importance metrics vary across the different machine learning models depicted. In AdaBoost, LightGBM, XGBoost, and CatBoost, feature importance is measured as the mean decrease in impurity or gain, indicating each feature’s contribution to reducing the overall prediction error. For SVM, feature importance is derived from the magnitude of the coefficients of the support vectors, reflecting the influence of features on the decision boundary. In DT, feature importance is determined by the reduction in impurity (Gini index or entropy) achieved by splitting the data based on each feature. ANN, KNN, and Naïve Bayes models use permutation importance, which quantifies the impact of feature perturbation on model performance. Consequently, these importance scores are on different scales and represent distinct notions of feature contribution, and therefore, direct comparisons across models should be made cautiously.

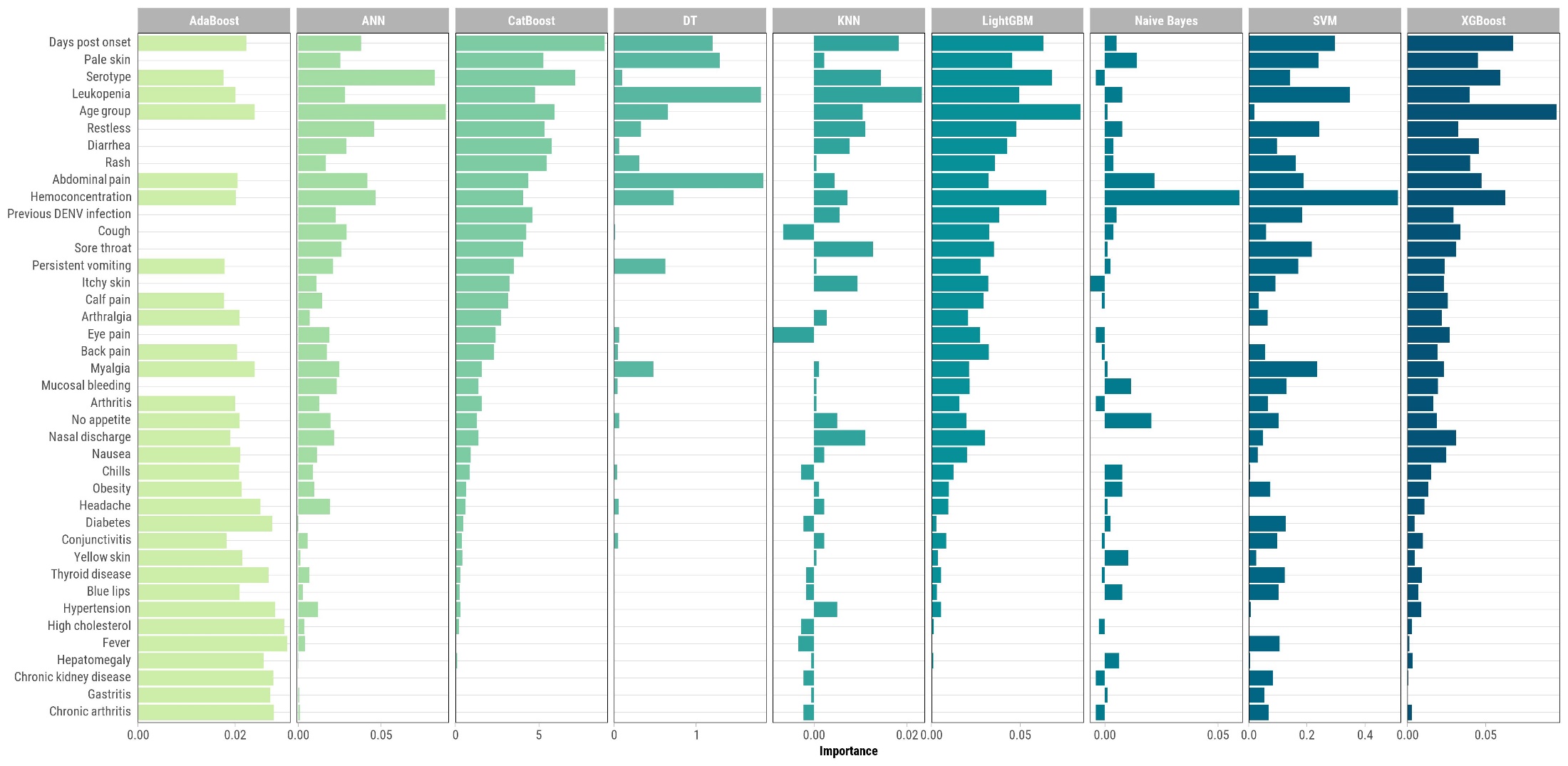

**Figure S4. Feature importance for each machine learning model for a reduced feature set which excludes leukopenia, hemoconcentration, DENV immune status, and DENV serotype, Sentinel Enhanced Dengue Surveillance System, Puerto Rico, 2012–2024**. The feature importance metrics vary across the different machine learning models depicted. In AdaBoost, LightGBM, XGBoost, and CatBoost, feature importance is measured as the mean decrease in impurity or gain, indicating each feature’s contribution to reducing the overall prediction error. For SVM, feature importance is derived from the magnitude of the coefficients of the support vectors, reflecting the influence of features on the decision boundary. In DT, feature importance is determined by the reduction in impurity (Gini index or entropy) achieved by splitting the data based on each feature. ANN, KNN, and Naïve Bayes models use permutation importance, which quantifies the impact of feature perturbation on model performance. Consequently, these importance scores are on different scales and represent distinct notions of feature contribution, and therefore, direct comparisons across models should be made cautiously.

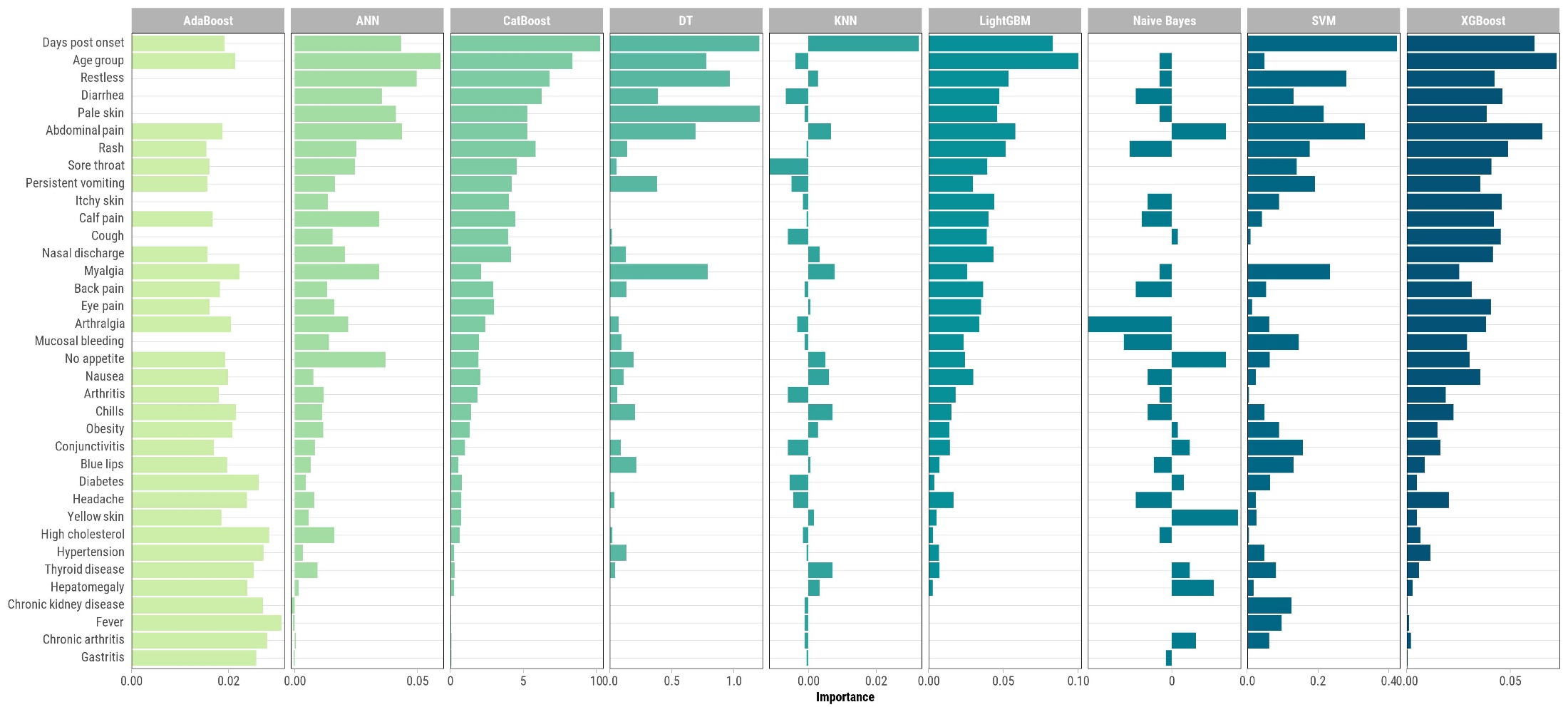
